# Integration of genomic classification and clinical characteristics predicts survival in metastatic prostate cancer

**DOI:** 10.64898/2026.01.12.26343673

**Authors:** Martin W. Schoen, Jiannong Li, Sihang Zeng, Heena Desai, Ryan Hausler, Candace L. Haroldsen, Lukas Owens, Luca F. Valle, Ruth B. Etzoni, Timothy R Rebbeck, Brent S. Rose, Michael J. Kelley, R. Bruce Montgomery, Nicholas G. Nickols, Matthew B. Rettig, Kosj Yamoah, Kara N. Maxwell, Isla P. Garraway

**Affiliations:** Saint Louis University School of Medicine, St. Louis MO; VA St Louis Healthcare System, St. Louis MO; Department of Biostatistics and Bioinformatics, H. Lee Moffitt Cancer Center, Tampa, Florida; James A. Haley Veterans’ Hospital, Tampa FL; Department of Biomedical Informatics and Medical Education, University of Washington, Seattle, WA, USA; Fred Hutchison Cancer Institute, Seattle, WA; Puget Sound VA Healthcare System, Seattle, WA; Medical Oncology Service, Corporal Michael Crescenz VA Medical Center, Philadelphia, Pennsylvania; Division of Hematology-Oncology, Department of Medicine, Perelman School of Medicine, University of Pennsylvania, Philadelphia; University of Utah, Salt Lake City, UT; Veterans Affairs Salt Lake City Healthcare System, Salt Lake City, UT; David Geffen School of Medicine at the University of California Los Angeles, Department of Radiation Oncology, Los Angeles, CA; UCLA Jonsson Comprehensive Cancer Center, Los Angeles, CA; Veterans Affairs Greater Los Angeles Healthcare System, Los Angeles, CA; Harvard T. H. Chan School of Public Health, Department of Epidemiology, Boston, MA; Dana-Farber Cancer Institute, Division of Medical Oncology, Boston, MA; Veterans Affairs Boston Healthcare System, Boston, MA; Veterans Affairs San Diego Healthcare System, San Diego, CA; University of California, San Diego, Department of Radiation Oncology, San Diego, CA; National Oncology Program, Department of Veteran Affairs, Washington; Durham VA Medical Center, Durham, NC; Department of Medicine and Duke Cancer Institute, Duke University, Durham, NC; Department of Hematology/Oncology, University of Washington, Seattle, WA; David Geffen School of Medicine at the University of California Los Angeles, Department of Hematology and Oncology, Los Angeles, CA; Department of Genetics, Perelman School of Medicine, Philadelphia, PA; David Geffen School of Medicine at the University of California Los Angeles, Department of Urology

**Keywords:** metastatic prostate cancer, genomic, Veterans, prognosis, survival

## Abstract

**Purpose:** Tumor comprehensive genomic profiling (CGP) has revolutionized cancer care and identifies patients for biomarker-specific therapy. In metastatic hormone-sensitive prostate cancer (mHSPC), CGP is not currently prognostic and no DNA-based genomic classification exists that accounts for combinations of alterations. We developed a DNA-based CGP classification that is prognostic for overall survival (OS) and could inform treatment.

**Methods:** Retrospective cross-sectional study using multivariable models to develop a clinico-genomic prognostic risk classification in U.S. Veterans diagnosed with synchronous mHSPC. Primary outcome was overall survival (OS) from time of metastatic diagnosis.

**Results:** 7201 Veterans with metastatic prostate cancer who underwent CGP were identified. There were 2484 Veterans (median [IQR] age 72 [67-77] years) with synchronous mHSPC and tissue CGP, which were divided into training and testing datasets. 16 genes associated with survival were identified and favorable, intermediate, and unfavorable genomic prognostication groups were created based upon mortality risk to generate the STRATOS-P classification. In a multivariable model, classification into intermediate and unfavorable groups was associated with increased mortality relative to the favorable group (aHR 1.54 [95% CI 1.33-1.78]; aHR 2.37 [95% CI 1.97-2.485], respectively), demonstrating an average AUC of 0.83. In an external validation cohort of non-Veterans, intermediate and unfavorable classifications were associated with increased mortality (aHR 2.45 [95% CI 1.87-3.21]; aHR 4.37 [95% CI 3.06-6.22], respectively) with an AUC of 0.79. The intermediate and unfavorable genomic prognostication groups were also associated with increased mortality across multiple disease states including synchronous and metachronous diagnoses, castration-resistance, and analyte type.

**Conclusions:** In metastatic prostate cancer, tumor DNA genomic alterations are prognostic for OS. The STRATOS-P classification is a validated prognostic tool that has the potential to guide decision-making in mHSPC.

## Background

Tumor DNA alterations are fundamental to understanding cancer biology. Disruption of tumor suppressor genes, such as *TP53*, carries prognostic importance^1^ while alterations in homologous recombination repair deficiency genes predict response to specific therapies, such as PARP inhibitors.^2^ In breast cancer, RNA-based gene expression assays, such as the 21-gene recurrence score (OncoType DX), are prognostic for risk of recurrence^3^ and overall survival.^4^ These tests inform clinical management, including the use of chemotherapy in early breast cancer.^5^ Similar integrative approaches in prostate cancer using RNA expression combined with clinical nomograms can improve prognostication and are being tested as potential predictive biomarkers.^6,7^

Prostate cancer is the most common non-cutaneous cancer in males with increasing incidence of disease.^8,9^ While metastatic disease is frequently lethal, survival has improved over time^10^ and many men do not die from their disease.^11^ Given the heterogeneity in survival and increasing treatment options, stratifying patients with metastatic disease into higher and lower risk groups could help to tailor treatment based on prognosis. In localized prostate cancer, where there are clinico-pathologic risk-stratification systems for biochemical recurrence,^12,13^ gene expression-based assays have been developed classify risk of metastatic progression,^6,7^ and multimodal AI models to predict benefit of androgen-deprivation therapy.^14,15^ In metastatic disease, risk stratification relies primarily on disease volume and timing of the onset of metastatic disease (synchronous versus metachronous) to predict benefits of therapy.^16–19^ Recently, a 22-gene RNA expression score was found to be prognostic in metastatic disease and predicted benefit from docetaxel in *PTEN*-inactivated tumors.^20^ There are many genomic alterations in prostate cancer that inform prognosis and may be potentially actionable.^21–25^

A wealth of data is collected in modern cancer care that includes patient characteristics, disease features, imaging, and comprehensive genomic profiling (CGP), many of which have not been integrated or used to their full clinical potential. CGP is recommended by the American Society of Clinical Oncology (ASCO), the Veterans Health Administration (VHA), and other national guidelines for all patients with metastatic prostate cancer.^26,27^ It is common for CGP to include DNA and RNA sequencing of over 300 genes, markers of microsatellite instability (MSI), and immunohistochemical tests. These data provide significant information about disease biology and may contribute to prognostication and/or prediction of response to therapies with little or no additional cost or burden on patients or clinicians.

The VHA serves over 16,000 Veterans with metastatic prostate cancer, including more than 2,000 diagnosed with metastatic disease per year.^28^ As a part of the National Precision Oncology Program (NPOP), somatic DNA CGP has been made available for every Veteran with metastatic prostate cancer at no cost to the Veteran.^29,30^ With the comprehensive clinical data of the VHA, a unique opportunity to classify tumor CGP results into prognostic categories exists without the need for additional sample acquisition or tumor analysis. Using CGP data from 7201 Veterans with metastatic prostate cancer, we developed and validated a prognostic risk classification system for patients with synchronous metastatic hormone sensitive prostate cancer (mHSPC) and tested its performance in an independent cohort and multiple metastatic prostate cancer disease states and analytes.

## Methods

### VA Data and Patient Population

The VHA Informatics and Computing Infrastructure (VINCI) was used to access the Corporate Data Warehouse (CDW) and clinicopathologic data from the VA-MAPP (Multi-OMICs Analysis Platform for Prostate Cancer) repository. Somatic tissue CGP with a commercially available platform (FoundationOne CDx or FoundationOne Liquid CDx; Foundation Medicine) was analyzed within the RESOLVE (Rate Elements Skewing Outcomes Linked to Veteran Equity) study via a data use agreement from the National Precision Oncology Program (NPOP). Analyses were performed in accordance with the Declaration of Helsinki. A waiver of consent was approved and results are reported according to the Strengthening the Reporting of Observational Studies in Epidemiology (STROBE) guidelines.

Veterans diagnosed with metastatic prostate cancer from 2003-2024 were identified by a natural language processing (NLP) algorithm^28^ and determined to be synchronous (diagnosis of prostate cancer less then 12 months before metastasis) or metachronous (diagnosis of prostate cancer 12 months or more before metastasis) (**Supplemental Table 1, Supplemental Figure 1**). At the time of sample collection, tumors were determined to be hormone-sensitive if there was no evidence of castration-resistance by an NLP algorithm within 3 months after tumor sampling or castration-resistant (CRPC) if obtained within 3 months before or any time after castration-resistance^31^ (**Supplemental Table 1, Supplemental Figure 1A**). Samples were divided into CGP performed on tissue (prostate or metastasis) or plasma (liquid biopsy), yielding eight cohorts (**Supplemental Table 1, Supplemental Figure 1A**). The date of diagnosis of metastatic disease was the start of observation (index date). Categorical variables, including age and individual gene alterations that were selected for inclusion were summarized using frequencies and proportions. The primary outcome for classification was overall survival, determined from diagnosis of metastasis to death or censoring, which was October 4, 2024.

### MSK-IMPACT validation data

Patients with metastatic cancer and tumor sequencing with Memorial-Sloan Kettering (MSK-IMPACT)^32^ was obtained from zenodo.^33^ In MSK-IMPACT, synchronous metastatic disease was identified if date of surgery (biopsy) was one year or less before metastases. Patients with only unspecified or male genital metastases were excluded. High volume disease was determined if metastases included lung, pleura, liver, biliary tract, and CNS metastases. Time from metastasis to death was the primary outcome and patient characteristics were determined across MSK-IMPACT datasets.^34^

### Creation of genomic prognostication groups

The discovery cohort included Veterans with mHSPC and CGP (n=2,484) and was randomly split into training (n=1,738) and test (n=746) sets at a 2:1 ratio, with overall survival divided between the two groups (**Supplementary Figure 1B**). Oncogenic somatic tumor DNA alterations including short variants, copy number alterations, and rearrangement variant calls were identified from CGP data as previously described,^35^ and genes were selected if found at >2% frequency in either synchronous mHSPC prostate or metastatic tissue. Genes were selected for inclusion were significantly associated with OS with Benjamini-Hochberg^36^ adjusted p-values of <0.05 in the training set using a multivariable Cox proportional hazards model including age, prostate specific antigen (PSA), and Charlson comorbidity index (CCI).^37^

Unsupervised hierarchical clustering was performed on the genomic alterations that were associated with OS in the training set using Jaccard distance and the Ward.D2 linkage method. Based on clustering results and clinical relevance determined by expert review, genomic alterations were categorized into three genomic groups using hazard ratios of less than <1, >1-2, or >2 for inclusion. Veterans with multiple alterations were categorized into the highest risk group. Additional prediction methods including an elastic net Cox model, random survival forest, and gradient boosting were tested to assess improved concordance.

### Statistical Analyses

The Kaplan–Meier method was used to estimate survival for the three prognostication groups in the synchronous mHSPC discovery cohort and a sub-cohort of 1389 Veterans with synchronous mHSPC whose CGP was ordered within six months of metastatic diagnosis. Model performance for predicting OS from 1 to 5 years was evaluated by the time-dependent area AUC (tAUC) using the discovery cohort test set was determined as the average of tAUC including age, PSA, CCI, year of diagnosis, volume of disease, and genomic classification. Missing numeric values imputed with the mean value, and all numeric values are standardized.^38^ Model performance using MSK-IMPACT external dataset was determined as the average of tAUC from 1 to 5 years using age, volume of disease, and genomic classification. Model performance for predicting OS from 1 to 5 years was also evaluated by the time-dependent area AUC (tAUC) in seven other VA cohorts based on diagnosis, hormonal sensitivity and analyte status (**Supplementary Figure 1B**). A sensitivity analysis of patients with complete data available was conducted between the synchronous mHSPC discovery cohort training and test sets including tumor volume from existing analyses^39^ and PSA. Statistical analyses were conducted using R software version 4.2.0 (https://www.R-project.org) and Python v3.10_using the scikit-survival package.^40^

## Results

Demographics, comorbidities, and clinic-pathological features of the synchronous mHSPC discovery cohort (n=2484) are reported in **Table 1**. The median age (IQR) at diagnosis of synchronous mHSPC was 72 (66-77) years with 765 Veterans (31%) self-identified as Black. The median (IQR) CCI was 2 (0-4) and BMI was 28.0 (24.6-31.6). Median (IQR) PSA at diagnosis was 47.6 ng/dL (14.9-176) and year of diagnosis was 2021 (IQR 2019-2022). Tissue for analysis was obtained from the prostate in 1951/2484 (78.5%) and from a metastatic site in 533/2484 (21.5%) (**Supplemental Figure 1**). Sequencing was performed within 6 months of metastatic diagnosis in 1,389 (55.9%) of Veterans and at a median of 3.5 months (IQR 0.5-22.2). The median overall survival was 59.4 months (95% CI 54.3-64.4) and 39.0 months (95% CI 34.4-43.5) in Veterans sequenced within 6 months of diagnosis of metastasis.

**Table 1.**
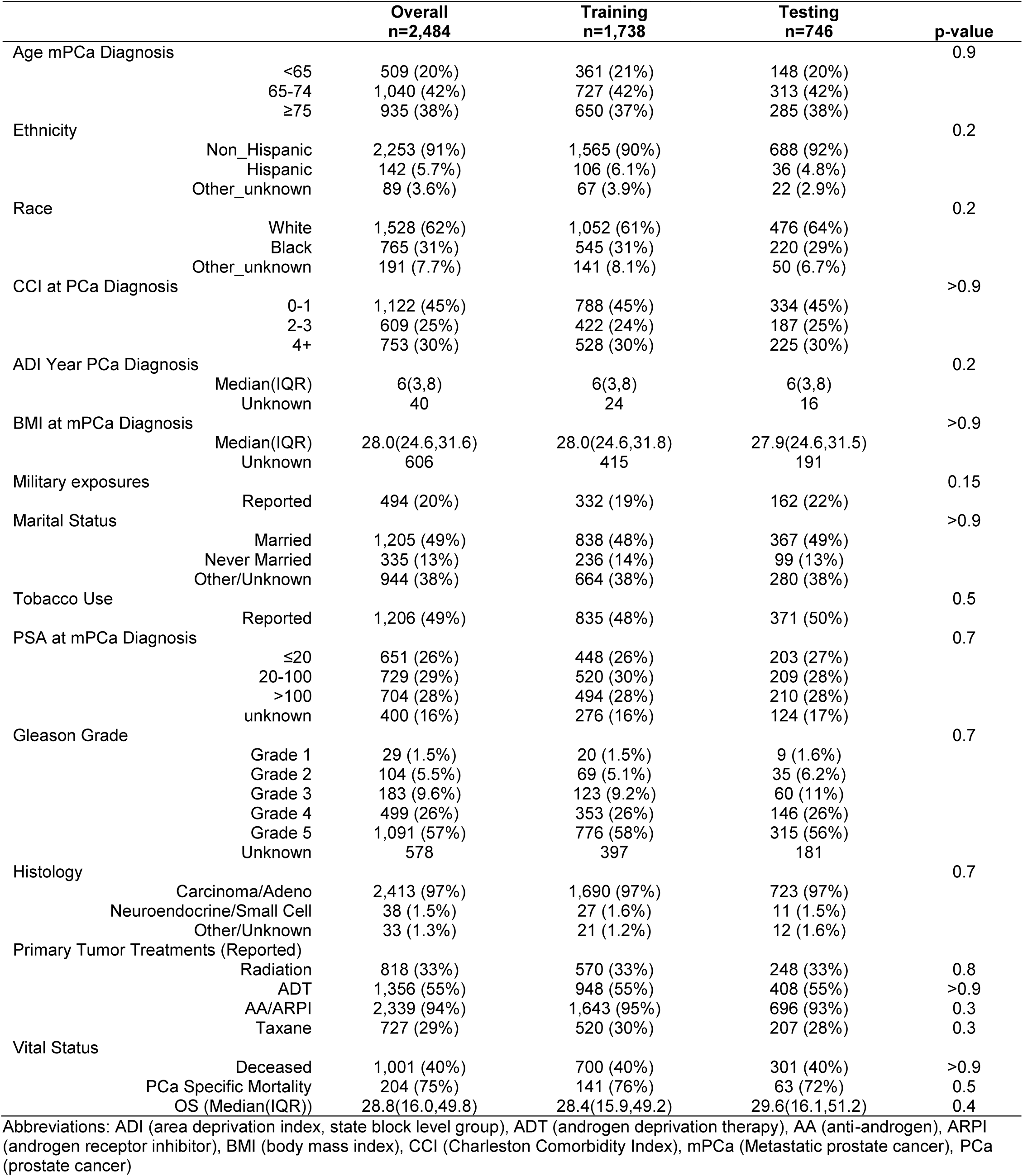
Patient characteristics of the synchronous metastatic HSPC Discovery Cohort. Table 1: Baseline characteristics of the discovery cohort of patients with synchronous metastatic hormone sensitive prostate cancer in all patients, the training subgroup, and the testing subgroup.

Results of CGP were used in the discovery cohort to identify pathogenic alterations prognostic for OS (**Figure 1A**). The association of alterations with OS was tested in a multivariable model and compared to samples with no alteration in that gene. After applying exclusion criteria, 16 genes were identified (**Figure 1A-B, Supplemental Table 2a-b**). These genes include the general tumor suppressor genes *TP53*, *PTEN*, and *RB1*, and genes involved in cell cycle regulation (*CCND1*, *CDK12*), DNA repair (*BRCA2*, *RAD21*), cell growth (*MYC, FGFR1, FGF3, FGF4, FGF19, PRKC1*, *LYN*), and androgen signaling (*AR, SPOP*). In the synchronous mHSPC discovery cohort, 70.1% (1741) of tumors sequenced had at least one oncogenic alteration, 28% (694) had two or more alterations, and 11.6% (289) had three or more (**Figure 1C**); 30% had no alteration in any of the 16 genes.

**Figure 1:**
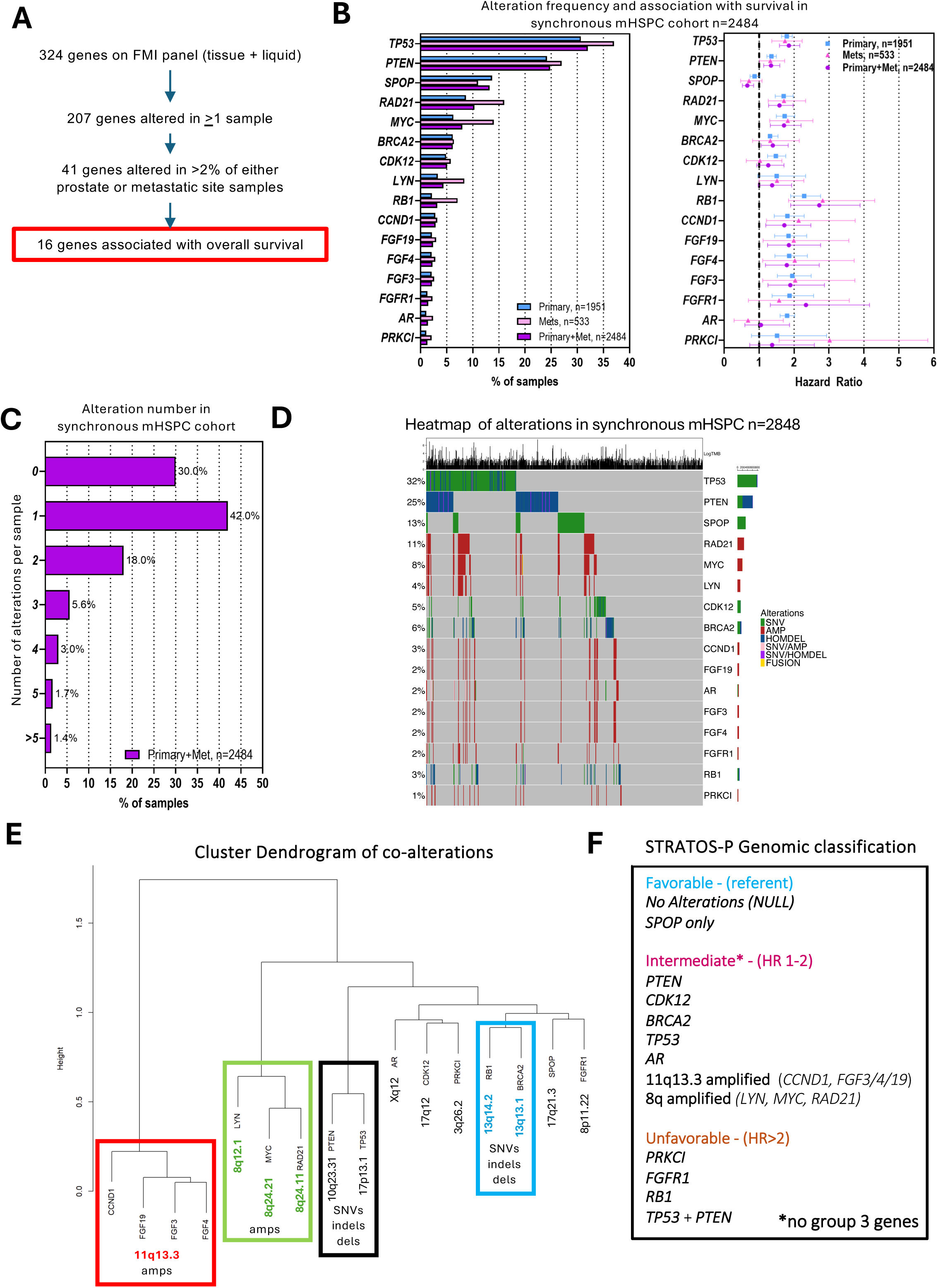
The incidence and types of alterations in patients in synchronous metastatic hormone sensitive prostate cancer. A, Selection of genes from Foundation Medicine Panel for inclusion in analysis. B, Incidence of genetic alterations based on site of biopsy in 16 genes selected and hazard ratio for overall survival for overall survival in multivariable model including age, Charlson Comorbidity Index. C, Distribution of the number of 16 gene alterations patients with synchronous metastatic hormone sensitive prostate cancer. D, Heatmap of alterations by gene and type synchronous metastatic hormone sensitive prostate cancer. E, Cluster dendrogram of alterations to display alterations that occur frequently together. F, Final grouping of DNA alterations in prognostic categories.

To appropriately classify genes, an analysis of co-alterations and mutual exclusivity was performed (**Figure 1D-E, Supplemental Table 2c)**. The most common co-alteration was *TP53* and *PTEN,* likely related to disease biology, as these genes are on different chromosomes (17p13.1 and 10q23.31, respectively). In contrast, other co-alterations likely occur due to commonly seen co-amplifications in prostate cancer, namely amplifications of 8q that involves *RAD21*, *LYN*, and *MYC* and 11q13.3, resulting in co-amplifications alterations of *CCND1, FGF19, FGF3*, and *FGF4*. Unsupervised hierarchical clustering of samples based on alterations confirmed these groupings (**Figure 1E)**.

Clustering, genomic co-localization, and hazard ratios from the multivariable model were used to group genes (**Figure 1F**). Genes with a HR of <1 or no alteration were grouped in the favorable risk category. One or more of the genes with HR of 1-2 and the presence of co-alterations were considered for inclusion into the intermediate risk group. Finally, the unfavorable risk group was created with genes with HR>2, and included the frequent co-alteration of *TP53* and *PTEN*, which was present in 9.5% (234/2484). Veterans were classified in to the most adverse group in which they had an alteration. In the synchronous mHSPC discovery cohort, 947/2484 (38%) of Veterans were classified as favorable risk while 1,180/2484 (48%) and 357/2484 (14%) were classified as intermediate and unfavorable risk, respectively (**Figure 2A**). Additional methods of classification showed no improvements in AUC, therefore the Cox model-based selection was used (**Supplemental Table 3**). Unadjusted overall survival of the discovery cohort was significantly lower in the intermediate (median OS 52 months, p<0.001) and unfavorable (median OS 31 months, p<0.001) groups compared to the favorable group (median OS 87 months) (**Figure 2B**). In Veterans with sequencing less than six months after diagnosis, unadjusted OS remained significantly lower in the intermediate (median OS 36 months, p<0.001) and unfavorable (median OS 24 months, p<0.001) groups compared to the favorable group (median OS not reached) (**Figure 2C**).

**Figure 2:**
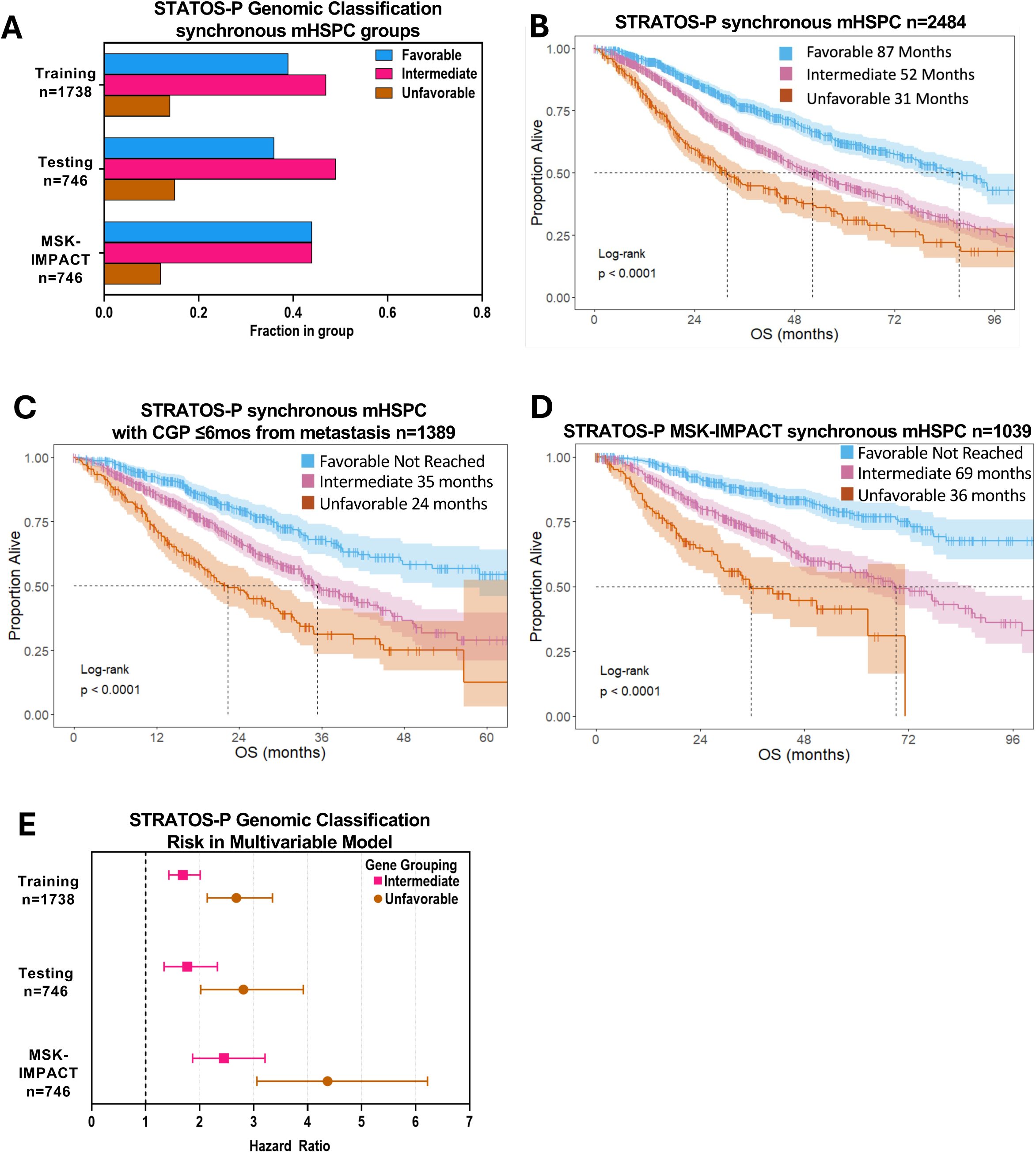
Distribution, overall survival and risk of mortality based on genomic classification. A, Frequency of genomic risk classification in synchronous metastatic hormone sensitive prostate cancer (mHSPC) in Veterans and Memorial Sloan Kettering-IMPACT. B, Overall survival in all patients with synchronous mHSPC in the VHA based on STRATOS-P (Somatic Tumor Risk Assessment for Overall Survival-Prostate) genomic classification. C, Overall survival in all patients with synchronous mHSPC who has genomic sequencing ordered within 6 months of diagnosis in the VHA based on the STRATOS-P genomic classification. D, Overall survival in patients identified with synchronous mHSPC in MSK IMPACT based on STRATOS-P genomic classification. E, Adjusted hazard ratio for death in synchronous mHSPC training, testing cohorts and MSK-IMPACT based on STRATOS-P genomic classification in the multivariable model.

The genomic classification was evaluated in the training dataset of 1,738 synchronous mHSPC Veterans to determine its performance in the multivariable model with age and CCI. In these Veterans, the adjusted hazard ratio (aHR) for mortality was 1.55 (95% CI 1.30-1.84, p<0.001) and 2.30 (95% CI 1.83-2.88, p<0.001) in the intermediate and unfavorable groups, respectively. In the test set of 746 Veterans held out from the discovery cohort of synchronous mHSPC, the aHR for mortality was 1.59 (95% CI 1.20-2.11) and 2.73 (95% CI 1.95-3.82) in the intermediate and unfavorable groups (**Figure 2C**, **Table 2**), respectively. The average tAUC was 0.83 using the full Cox model.

**Table 2:** Association of genomic prognostication groups with overall survival in multivariable Cox model. Table 2: Multivariable Cox model features and for patients with synchronous metastatic hormone sensitive prostate cancer based on genomic classification, age, Charlson Comorbidity Index (CCI) and tumor volume that was available. Results are shown for the training subgroup, the testing subgroup, and with MSK-IMPACT. Time-dependent and average AUC measurement between the training subgroup and both the testing subgroup and MSK-IMPACT with volume of disease and genomic classification are shown.

|  | Training, n=1738 |  | Testing, n=746 |  | MSK IMPACT, n=1039 |  |
| --- | --- | --- | --- | --- | --- | --- |
|  | n (%) | HR (95% CI) | n (%) | HR (95% CI) | n (%) | HR (95% CI) |
| Age <65 | 361 (21%) | Ref | 148 (20%) | Ref | 492 (47%) | Ref |
| Age 65-74 | 727 (42%) | 1.18 (0.96, 1.45) | 313 (42%) | 1.46 (1.04, 2.06) | 382 (37%) | 1.29 (0.99, 1.68) |
| Age >75 | 650 (37%) | 1.40 (1.12, 1.74) | 285 (38%) | 1.63 (1.13, 2.33) | 165 (16%) | 2.61 (1.93, 3.53) |
| CCI 0-1 | 788 (45%) | Ref | 334 (45%) | Ref | NA |  |
| CCI 2-3 | 422 (24%) | 1.35 (1.12, 1.65) | 187 (25%) | 1.02 (0.75, 1.39) | NA |  |
| CCI 4+ | 528 (30%) | 1.71 (1.43, 2.03) | 225 (30%) | 1.80 (1.37, 2.36) | NA |  |
| PSA <20 | 448 | ref | 203 | ref | NA |  |
| PSA 20-100 | 520 | 1.05 (0.86, 1.29) | 209 | 1.04 (0.76, 1.42) | NA |  |
| PSA>100 | 494 | 1.38 (1.13, 1.68) | 210 | 1.41 (1.04, 1.92) |  |  |
| Unknown | 276 | 0.71 (0.50, 0.99) | 124 | 0.79 (0.47, 1.34) | NA |  |
| Low volume | 401 (23%) | Ref | 177 (24%) | Ref | 782 (75.3%) | Ref |
| High volume | 605 (35%) | 1.53 (1.25, 1.87) | 270 (36%) | 2.31 (1.66, 3.3) | 257 (24.7%) | 3.78 (3.00, 4.76) |
| Unknown | 732 (42%) | 0.84 (0.68, 1.05) | 299 (40%) | 1.28 (0.89, 1.83) | 0 (0) |  |
| Diagnosis year |  |  |  | 1.27 (1.18, 1.37) |  |  |
| Favorable | 681 (39%) | Ref | 266 (36%) | Ref | 457 | Ref |
| Intermediate | 815 (47%) | 1.65 (1.39, 1.97) | 365 (49%) | 1.68 (1.27, 2.24) | 453 | 2.19 (1.67, 2.88) |
| Unfavorable | 242 (14%) | 2.44 (1.95, 3.07) | 115 (15%) | 2.99 (2.12, 4.20) | 129 | 3.92 (3.00, 4.75) |
| **AUC** |  |  | **Testing, n=746** |  | **MSK IMPACT, n=1014** |  |
| **time (months)** |  |  | **Age+CCI** | **Full Model\*** | **Age** | **Full Model#** |
| 12 |  |  | 0.69 | 0.82 | 0.65 | 0.81 |
| 24 |  |  | 0.69 | 0.80 | 0.64 | 0.79 |
| 36 |  |  | 0.71 | 0.83 | 0.63 | 0.78 |
| 48 |  |  | 0.73 | 0.88 | 0.65 | 0.78 |
| 60 |  |  | 0.72 | 0.88 | 0.64 | 0.78 |
| **Average AUC** |  |  | **0.71** | **0.83** | **0.64** | **0.79** |

NA: Not Available, AUC: Area Under the receiver operating characteristic Curve
\* Included age, CCI, PSA, volume of disease, year of diagnosis, and genomic classification

In the MSK-IMPACT validation cohort of 1039 patients with synchronous mHSPC (**Supplemental Table 4**) the median survival was 89.8 months (95% CI 72.1-107.4) (**Figure 2D**). The favorable group included 44.0% (457/1039) patients, the intermediate group included 43.6% (453/1039), and the unfavorable group 12.4% (129/1039) (**Figure 2A**). Using age and genomic classification, the aHR for mortality in MSK-IMPACT of the intermediate group was 2.45 (95% CI 1.87-3.21) and 4.37 (95% CI 3.06-6.22) in the unfavorable group (**Table 2**, **Figure 2E**). The average tAUC over 12-60 months was 0.79 between the discovery cohort and MSK-IMPACT (**Table 2**).

In 1389 Veterans who had CGP within 6 months, mortality was slightly higher compared to the overall cohort with an aHR of 1.90 (95% CI 1.52-2.38) and 3.16 (95% CI 2.44-4.11) in the intermediate and unfavorable groups, respectively. The tAUC was 0.72 at 12 months (0.68-0.75) in the test set in those with sequencing within 6 months. Additional sensitivity analyses in veterans with known tumor volume (n=787), PSA at diagnosis (n=1462), or both (n=733). Adding tumor volume improved the tAUC at 12 months to 0.79, and adding PSA did not change the tAUC at 12 months, also 0.77 (**Supplemental Table 5**).

The genomic classifier was assessed across other chronologic metastatic presentations, hormonal sensitivity, and sequencing analytes (**Supplemental Table 1**). In 384 Veterans with synchronous mHSPC who had a liquid biopsy, mortality was similar to the tissue analyte with an aHR of 2.23 (95% CI 1.45, 3.43) and 5.13 (95% CI 2.93-8.97) in the intermediate and unfavorable groups, respectively, and the tAUC was 0.71 at 12 months (0.71-0.77) (**Figure 3, Supplemental Table 6**). In Veterans diagnosed with synchronous metastases but CGP was obtained during castration-resistance (tissue, n=336); liquid biopsy (n=691), the genomic classification remained associated with OS with acceptable performance in tissue [tAUC 0.70 (0.69-0.73)] and in liquid biopsy [tAUC 0.74 (0.70-0.78)] at 12 months. In Veterans with a metachronous mHSPC sample, the genomic classification was associated with mortality when either tissue (n=1236) or liquid (n=297) was analyzed [tAUC 0.70 (0.63-0.77) and 0.79 (0.75-0.84), respectively]. In Veterans with a metachronous mCRPC sample, the genomic classification was associated with mortality when either tissue (n=488) or liquid (n=1285) was analyzed [tAUC 0.70 (0.63-0.77) and 0.72 (0.68-0.78), respectively].

**Figure 3:**
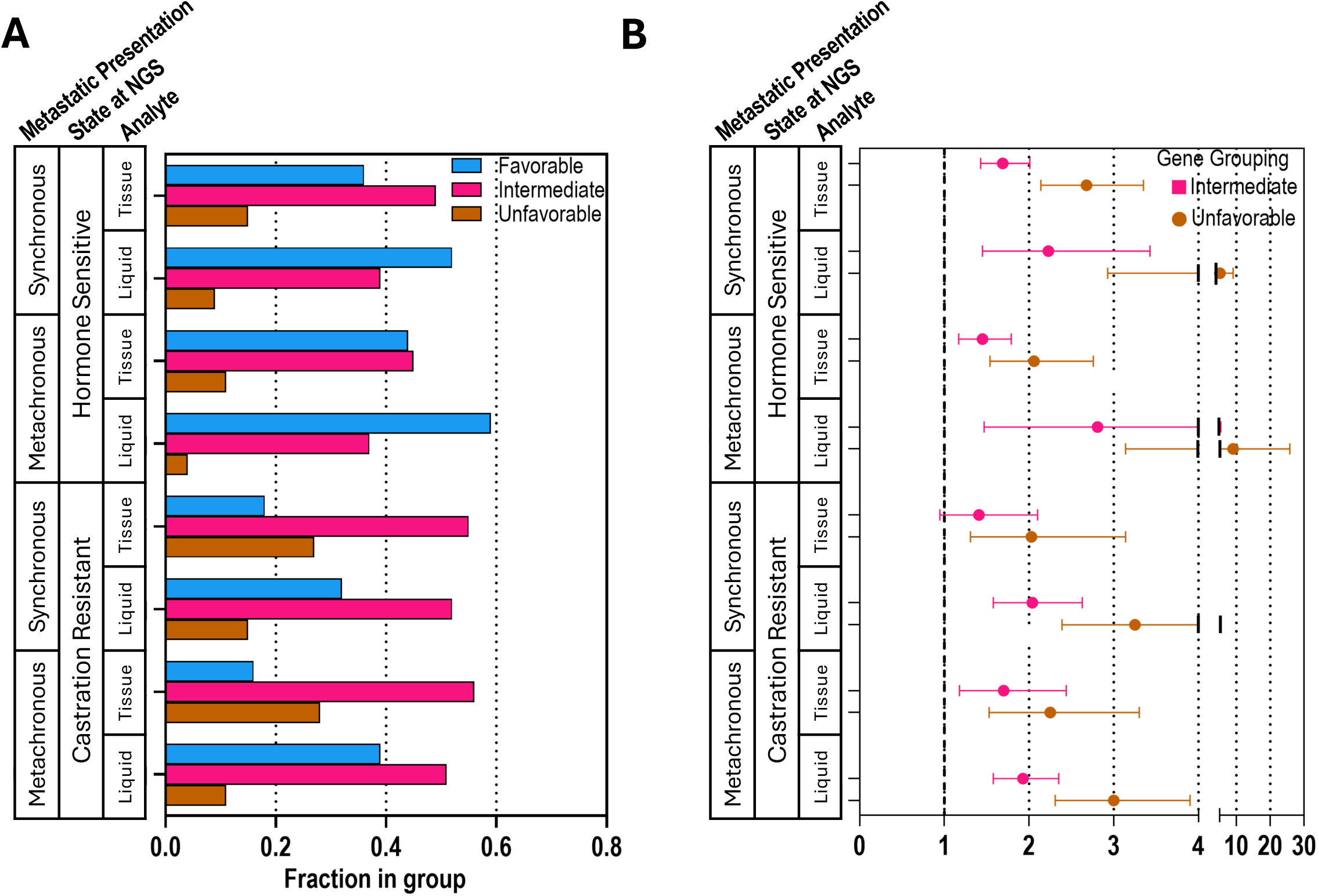
Analyses from VHA validation cohorts. A, Incidence of alteration groups based on stage at biopsy, tissue vs. liquid, and synchronous vs. metachronous presentation B, Adjusted hazard ratio for validation cohorts based in disease state at time of sequencing, tissue vs. liquid biopsy, synchronous or metachronous, and type of biopsy.

## Discussion

This large, multicenter study demonstrates that a genomic classification of somatic DNA CGP provides powerful, reproducible prognostic information in metastatic prostate cancer. To date, CGP is not prognostic in synchronous mHSPC as there is no validated classification system. Our genomic classification, termed STRATOS-P (Somatic Tumor Risk Assessment for Overall Survival-Prostate), differentiates risk using a large commercially available panel. The use of existing CGP for prognosis without additional testing helps to understand heterogeneity in survival outcomes and has the potential to influence therapeutic decisions and more precisely tailor clinical trials for escalation/de-escalation strategies.

STRATOS-P was trained and validated in veterans with synchronous mHSPC and non-Veterans with synchronous mHSPC from MSK-IMPACT. Sensitivity analyses restricting to synchronous mHSPC with CGP within 6 months of diagnosis to account for immortal time bias showed similar results. The performance was similar when applied to patients with metachronous mHSPC and when CGP was performed on tissue in the mCRPC clinical state, demonstrating the robustness of STRATOS-P in different clinical scenarios. To date, no other clinical variable had as much ability to differentiate risk of mortality in various disease states, underscoring the importance of DNA genomic alterations in prognosis. It is important that STRATOS-P performed well in liquid biopsies because tumor tissue may be either unavailable or difficult to obtain.

Given the increase in metastatic prostate cancer incidence and treatment options, it is important to consider the heterogeneity of these tumors.^8^ The combination of clinical and genomic features contribute to the promise of precision oncology to optimize treatment that could prolong survival, while considering quality of life and adverse events. As prostate cancer is a disease of aging, estimation of prognosis is especially important for frail patients or those at risk for cardiac events to avoid over-treatment. In frail patients with low-risk metastatic prostate cancer, combination therapies may have little effect on mortality and may increase adverse events, as has been shown in trials of fit patients.^41^

### Limitations and Strengths

Because of its retrospective design, this study is subject to unmeasured confounding. Veterans who received CGP may have different characteristics from other patients with mHSPC, biasing the results. Furthermore, because CGP historically only affected treatment decisions for mCRPC, many patients had CGP several months after initial diagnosis. This immortal time and left truncation biases was addressed through an analysis of patients sequenced within six months of diagnosis. Also, patients were identified as “metastatic” through a natural language processing (NLP) of VHA electronic health records. While accurate, it includes a population of patients with both regional metastases and distant metastasis.^28^ Lastly, veterans may have received treatment outside of the VA Healthcare System, creating uncertainty in estimates of the timing/accuracy of synchronous diagnosis and comorbid conditions.

The study has several strengths, most notably, its large sample size. The ability to perform discovery and validation of genomic alterations in an initial cohort of over 7000 patients is important for creating a comprehensive classification. The large number of samples allows for management of co-alterations and an understanding of the interactions that commonly occur in tumors. The validation of the classification in other VA data and the external MSK-IMPACT dataset increases the reliability of our findings. The use of existing CGP results without additional sampling or cost will also increase the ability to apply these findings in practice and clinical trials.

### Conclusion

The STATOS-P genomic classification using DNA alterations from CGP is prognostic and was validated in internal VA data and the external non-Veteran MSK-IMPACT cohort. This classification does not require additional tissue procurement or sequencing since it uses existing data obtained as part of contemporary clinical practice. In combination with clinical variables, STRATOS-P categorizes patients with prostate cancer into distinct prognostic risk groups. These groups could be used to inform shared decision-making to inform treatment options and for prospective studies to evaluate disease risk combined with clinical variables to select and optimize treatment.

## Data Availability

All data produced in the present study are available upon reasonable request to the authors and subject to a data use agreement

## Author Contributions

Dr. Schoen, Maxwell, and Garraway had full access to all of the data in the study and take responsibility for the integrity of the data and the accuracy of the data analysis.

### Concept and design

Garraway, Maxwell, Schoen, Yamoah, Nickols, Rebbeck, Rettig.

### Acquisition, analysis, or interpretation of data

Schoen, Li, Zeng, Owens, Maxwell, Garraway, Rettig, Kelley.

### Drafting of the manuscript

Schoen, Maxwell, Garraway.

### Critical review of the manuscript for important intellectual content

Schoen, Garraway, Maxwell, Valle, Owens, Zeng, Etzioni, Rebbeck, Nichols, Montgomery, Rettig, Yamoah, Kelley

### Statistical analysis

Schoen, Li, Zeng, Owens, Desai, Hausler, Haroldsen.

### Obtained funding

Schoen, Maxwell, Garraway, Yamoah, Rebbeck, Rose, Montgomery.

### Administrative, technical, or material support

Hausler, Desai, Haroldsen.

### Supervision

Schoen, Maxwell, Garraway.

## Disclaimer

The views expressed are those of the authors and do not necessarily reflect the position or policy of the US Department of Veterans Affairs or the US government.

---

**Supplementary Fig 1:**
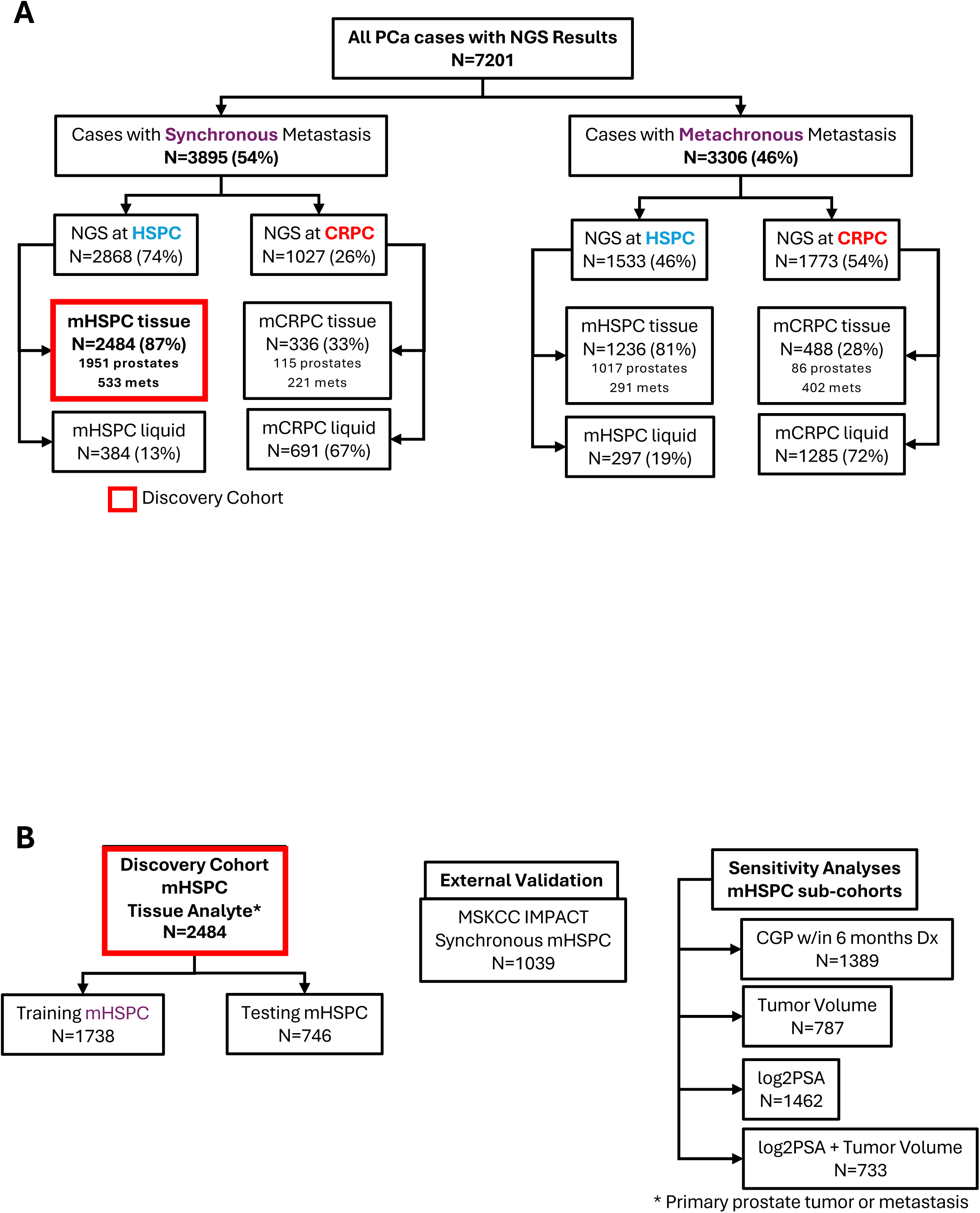
Cohort creation for genomic classification. A, Complete categorization of all samples of patients with metastatic prostate cancer from National Precision Oncology Program (NPOP). The date of sample acquisition was used to determine the state of disease. Patients with no evidence of prostate cancer prior to 12 months before metastatic diagnosis were considered to be synchronous while patients with a diagnosis of prostate cancer prior to 12 months were considered to be metachronous. Samples acquired from patients within 90 days prior to castration resistance were considered castration-resistant while samples acquired more than 90 days prior to castration-resistance were considered hormone sensitive. Tissue samples that were determined to be synchronous metastatic hormone sensitive prostate cancer (mHSPC) were used to create the discovery cohort to initially classify genomic alterations. B, Creation of the training and testing sub-groups of patients with mHSPC in the discovery cohort and the primary validation cohorts of MSK-IMPACT and sensitivity analyses in specific cohorts of patients with synchronous mHSPC.

**Supplementary Fig 2:**
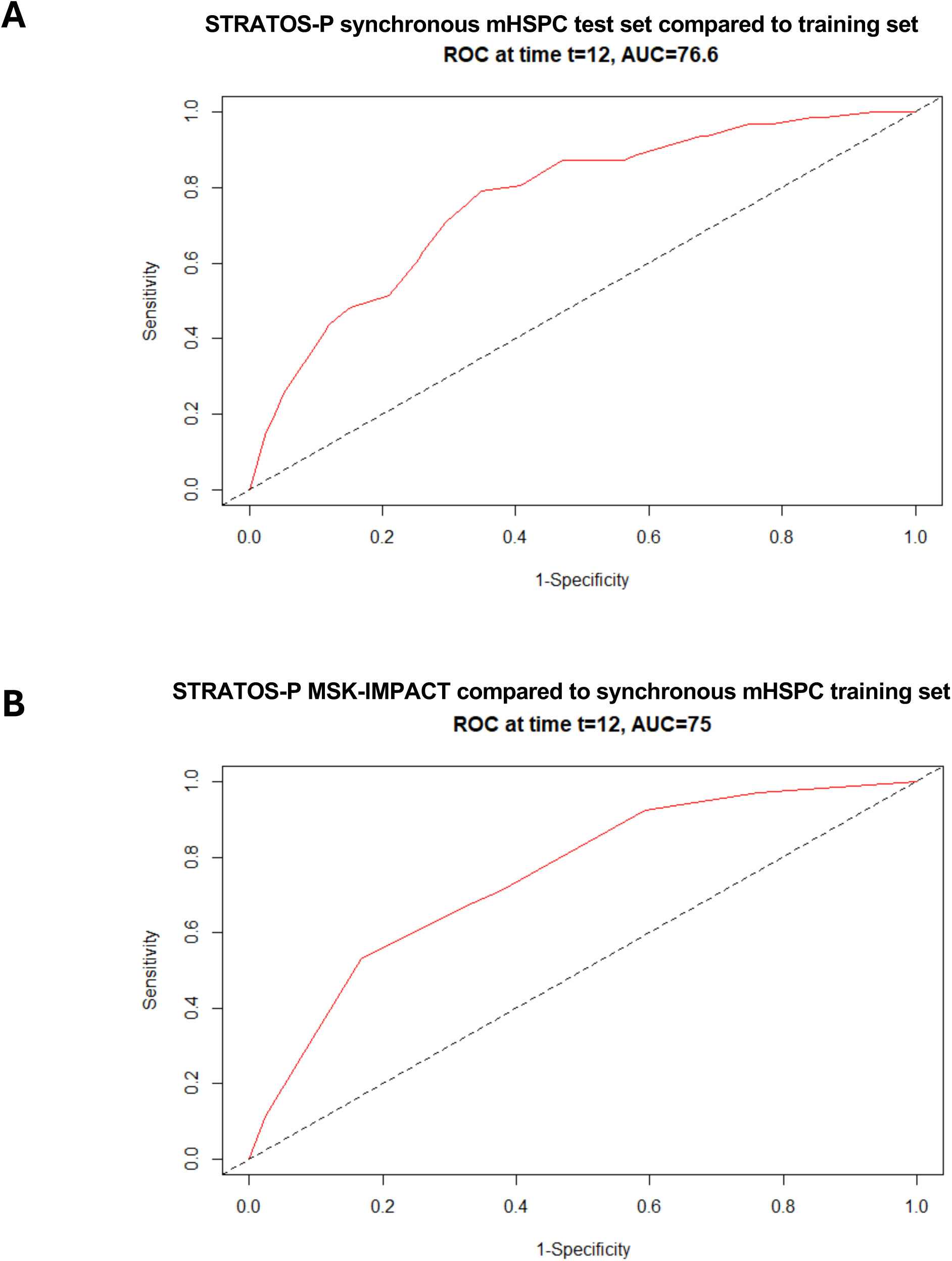
Receiver operating characteristic (ROC) curves for STRATOS-P. A, ROC for synchronous mHSPC test set compared to the synchronous mHSPC training set. B, ROC for MSK-IMPACT compared to the synchronous mHSPC training set.

**Supplementary Table 1:** Cohort definitions and numbers.

| Metastatic presentation (1) | Sequenced Tissue Type (2) | Sequencing State (3) | Description in words | n in Cohort (total n=7201) | % of Cohort (total n=7201) | # of prostates | # of metastases |
| --- | --- | --- | --- | --- | --- | --- | --- |
| Synchronous | Tissue | HSPC | Synchronous/Denovo, HSPC when sequenced tissue | 2484 | 34.5% | 1951 | 533 |
| Synchronous | Liquid | HSPC | Synchronous/Denovo, HSPC when sequenced liquid | 384 | 5.3% | n/a | n/a |
| Synchronous | Tissue | CRPC | Synchronous/Denovo, CRPC when sequenced tissue | 336 | 4.7% | 115 | 221 |
| Synchronous | Liquid | CRPC | Synchronous/Denovo, CRPC when sequenced liquid | 691 | 9.6% | n/a | n/a |
| Metachronous | Tissue | HSPC | Metachronous/recurrent, HSPC when sequenced tissue | 1236 | 17.2% | 1017 | 219 |
| Metachronous | Liquid | HSPC | Metachronous/recurrent, HSPC when sequenced liquid | 297 | 4.1% | n/a | n/a |
| Metachronous | Tissue | CRPC | Metachronous/recurrent, CRPC when sequenced tissue | 488 | 6.8% | 86 | 402 |
| Metachronous | Liquid | CRPC | Metachronous/Recurrent, CRPC when sequenced liquid | 1285 | 17.8% | n/a | n/a |

(1) Metastatic presentation is defined as synchronous = metastatic diagnosis date less than 12 months from prostate cancer diagnosis date or metachronous = metastatic diagnosis date at least 12 months after prostate cancer diagnosis date
(2) Sequenced Tissue Type is defined as Tissue = FoundationOneCdx testing on a prostate or metastasis from prostate cancer OR Liquid = FoundationOneCdx testing on cell-free DNA (liquid biopsy) from prostate cancer patient
(3) Sequencing State is defined as Hormone sensitive prostate cancer (HSPC) = no diagnosis of castrate resistant prostate cancer (CRPC) or CRPC date is >90 days after specimen collection date OR CRPC = CRPC date is prior to OR <90 days after specimen collection date

**Supplementary Table 2a:** Association of alterations in gene with overall survival identified 16 genes.

| Gene | n | % of 2484 | Primary+Met (n=2484) |  |  | n | % of 1951 | Primary only (n=1951) |  |  | n | % of 533 | Met only (n=533) |  |  |
| --- | --- | --- | --- | --- | --- | --- | --- | --- | --- | --- | --- | --- | --- | --- | --- |
|  |  |  | HR (95% CI) | P (Wald) | adjusted p BH |  |  | HR (95% CI) | P (Wald) | adjusted p BH |  |  | HR (95% CI) | P (Wald) | adjusted p BH |
| AR | 34 | 1.4% | 1.05 (0.59, 1.87) | 8.55E-01 | 9.91E-01 | 21 | 1.1% | 1.80 (1.60, 2.02) | 6.11E-23 | 6.89E-21 | 13 | 2.4% | 0.68 (0.28, 1.69) | 4.10E-01 | 9.98E-01 |
| BRCA2 | 154 | 6.2% | 1.39 (1.06, 1.83) | 1.75E-02 | 1.41E-01 | 120 | 6.2% | 1.31 (1.11, 1.54) | 1.41E-03 | 1.53E-02 | 34 | 6.4% | 1.32 (0.81, 2.15) | 2.66E-01 | 8.43E-01 |
| CCND1 | 72 | 2.9% | 1.72 (1.19, 2.48) | 3.86E-03 | 4.49E-02 | 55 | 2.8% | 1.81 (1.43, 2.29) | 6.74E-07 | 1.63E-05 | 17 | 3.2% | 2.13 (1.21, 3.75) | 8.51E-03 | 1.00E-01 |
| CDK12 | 126 | 5.1% | 1.26 (0.93, 1.71) | 1.29E-01 | 6.02E-01 | 95 | 4.9% | 1.48 (1.24, 1.76) | 1.66E-05 | 3.11E-04 | 31 | 5.8% | 1.02 (0.63, 1.65) | 9.29E-01 | 9.98E-01 |
| FGF19 | 59 | 2.4% | 1.85 (1.24, 2.76) | 2.66E-03 | 3.97E-02 | 43 | 2.2% | 1.84 (1.44, 2.36) | 1.22E-06 | 2.57E-05 | 16 | 3.0% | 1.99 (1.11, 3.57) | 2.11E-02 | 2.08E-01 |
| FGF3 | 55 | 2.2% | 1.89 (1.25, 2.87) | 2.73E-03 | 3.97E-02 | 41 | 2.1% | 1.95 (1.52, 2.49) | 1.20E-07 | 3.39E-06 | 14 | 2.6% | 2.03 (1.10, 3.74) | 2.28E-02 | 2.17E-01 |
| FGF4 | 56 | 2.3% | 1.79 (1.18, 2.72) | 6.02E-03 | 6.64E-02 | 41 | 2.1% | 1.86 (1.45, 2.38) | 1.07E-06 | 2.41E-05 | 15 | 2.8% | 2.02 (1.10, 3.72) | 2.40E-02 | 2.20E-01 |
| FGFR1 | 37 | 1.5% | 2.34 (1.32, 4.16) | 3.68E-03 | 4.49E-02 | 25 | 1.3% | 1.87 (1.37, 2.56) | 9.49E-05 | 1.60E-03 | 12 | 2.3% | 1.57 (0.69, 3.58) | 2.80E-01 | 8.43E-01 |
| LYN | 109 | 4.4% | 1.37 (0.97, 1.93) | 7.16E-02 | 4.09E-01 | 64 | 3.3% | 1.50 (0.96, 2.33) | 7.61E-02 | 4.13E-01 | 45 | 8.4% | 1.51 (1.00, 2.28) | 4.83E-02 | 3.20E-01 |
| MYC | 199 | 8.0% | 1.71 (1.32, 2.20) | 3.82E-05 | 1.59E-03 | 122 | 6.3% | 1.73 (1.49, 2.02) | 2.02E-12 | 1.14E-10 | 77 | 14.0% | 1.82 (1.30, 2.54) | 4.55E-04 | 1.25E-02 |
| PRKCI | 33 | 1.3% | 1.37 (0.73, 2.58) | 3.22E-01 | 8.63E-01 | 22 | 1.1% | 1.51 (0.78, 2.93) | 2.24E-01 | 7.37E-01 | 11 | 2.1% | 3.02 (1.57, 5.83) | 9.59E-04 | 1.97E-02 |
| PTEN | 616 | 24.8% | 1.34 (1.13, 1.59) | 5.93E-04 | 1.44E-02 | 472 | 24.2% | 1.35 (1.22, 1.49) | 5.34E-09 | 1.85E-07 | 144 | 27.0% | 1.32 (1.01, 1.73) | 4.30E-02 | 2.95E-01 |
| RAD21 | 257 | 10.3% | 1.58 (1.27, 1.97) | 3.65E-05 | 1.59E-03 | 170 | 8.7% | 1.70 (1.46, 1.97) | 4.59E-12 | 2.21E-10 | 87 | 16.0% | 1.71 (1.26, 2.33) | 6.13E-04 | 1.52E-02 |
| RB1 | 80 | 3.2% | 2.72 (1.90, 3.89) | 3.91E-08 | 3.79E-06 | 42 | 2.2% | 2.29 (1.90, 2.76) | 6.88E-18 | 5.82E-16 | 38 | 7.1% | 2.82 (1.84, 4.31) | 1.74E-06 | 2.14E-04 |
| SPOP | 328 | 13.2% | 0.66 (0.52, 0.84) | 7.83E-04 | 1.75E-02 | 267 | 13.7% | 0.87 (0.74, 1.02) | 7.86E-02 | 2.86E-01 | 61 | 11.0% | 0.71 (0.47, 1.08) | 1.11E-01 | 5.27E-01 |
| TP53 | 794 | 32.0% | 1.85 (1.58, 2.16) | 5.30E-15 | 1.54E-12 | 599 | 30.7% | 1.79 (1.64, 1.95) | 3.98E-38 | 1.34E-35 | 195 | 37.0% | 1.74 (1.36, 2.23) | 1.26E-05 | 1.03E-03 |

**Supplementary Table 2b:** Published genomic chracteristics of 16 genes selected.

| Gene | Chr:Pos | Band | % of<br>2484<br>dmHSPC | % of 1951<br>dmHSPC<br>prostate | % of 533<br>dmHSPC<br>metastasis | Overall<br>Alteration<br>Freq cbio | Mutation<br>Freq cbio | CNA Freq<br>cbio | CNA Type | Struc Rearr<br>Freq cbio |
| --- | --- | --- | --- | --- | --- | --- | --- | --- | --- | --- |
| *AR* | chrX:67,544,021-67,730,619 | Xq12 | 1.4% | 1.1% | 2.4% | 14.6% | 4.1% | 10.5% | amp | <1% |
| *BRCA2* | chr13:32,315,508-32,400,268 | 13q13.1 | 6.2% | 6.2% | 6.4% | 6.2% | 4.0% | 2.2% | homodel | <1% |
| *CCND1* | chr11:69,641,156-69,654,474 | 11q13.3 | 2.9% | 2.8% | 3.2% | 2.4% | <1% | 2.4% | amp | <1% |
| *CDK12* | chr17:39,461,486-39,534,544 | 17q12 | 5.1% | 4.9% | 5.8% | 5.8% | 5.8% | <1% | - | <1% |
| *FGF19* | chr11:69,698,238-69,704,022 | 11q13.3 | 2.4% | 2.2% | 3.0% | 2.0% | <1% | 2.0% | amp | <1% |
| *FGF3* | chr11:69,809,968-69,819,416 | 11q13.3 | 2.2% | 2.1% | 2.6% | 1.6% | <1% | 1.6% | amp | <1% |
| *FGF4* | chr11:69,771,022-69,775,341 | 11q13.3 | 2.3% | 2.1% | 2.8% | 1.7% | <1% | 1.7% | amp | <1% |
| *FGFR1* | chr8:38,411,143-38,468,635 | 8p11.22 | 1.5% | 1.3% | 2.3% | 3.2% | <1% | 1.7%/1.5% | amp/homodel | <1% |
| *LYN* | chr8:55,879,835-56,014,169 | 8q12.1 | 4.4% | 3.3% | 8.4% | 2.0% | <1% | 2.0% | amp | <1% |
| *MYC* | chr8:127,736,231-127,742,951 | 8q24.21 | 8.0% | 6.3% | 14.0% | 6.8% | 1.0% | 5.8% | amp | <1% |
| *PRKCI* | chr3:170,222,424-170,305,977 | 3q26.2 | 1.3% | 1.1% | 2.1% | 0.0% | <1% | <1% | - | <1% |
| *PTEN* | chr10:87,863,625-87,971,930 | 10q23.31 | 24.8% | 24.2% | 27.0% | 21.0% | 8.1% | 11.6% | homodel | 1.3% |
| *RAD21* | chr8:116,845,934-116,874,776 | 8q24.11 | 10.3% | 8.7% | 16.0% | 3.0% | <1% | 3.0% | amp | <1% |
| *RB1* | chr13:48,303,751-48,481,890 | 13q14.2 | 3.2% | 2.2% | 7.1% | 6.6% | 3.2% | 3.4% | homodel | <1% |
| *SPOP* | chr17:49,598,884-49,678,097 | 17q21.3 | 13.2% | 13.7% | 11.0% | 14.1% | 14.1% | - | - | <1% |
| *TP53* | chr17:7,668,421-7,687,490 | 17p13.1 | 32.0% | 30.7% | 37.0% | 32.1% | 28.8% | 2.2% | homodel | 1.1% |

**Supplementary Table 2c:**
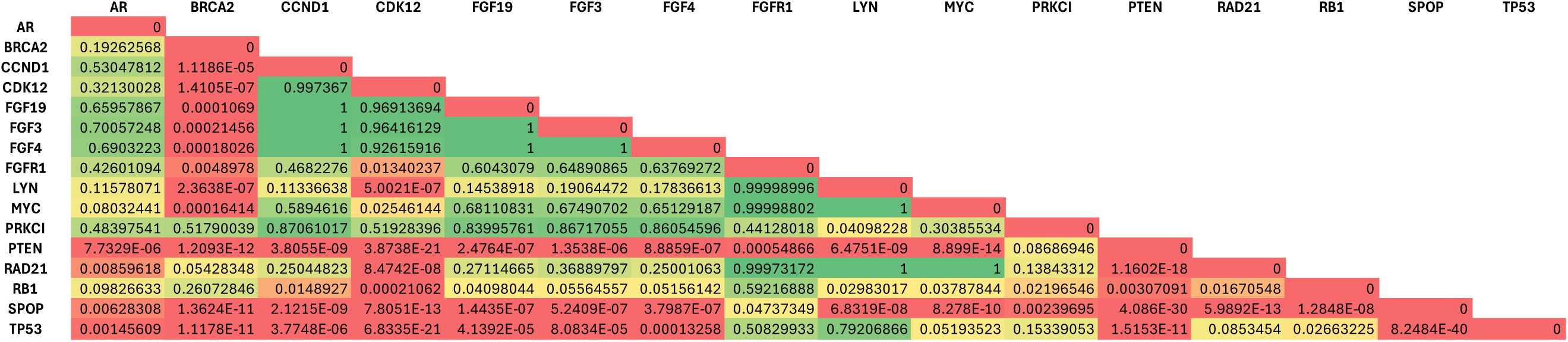
Mutual exclusivity of 16 genes selected.

|  | AR | BRCA2 | CCND1 | CDK12 | FGF19 | FGF3 | FGF4 | FGFR1 | LYN | MYC | PRKCI | PTEN | RAD21 | RB1 | SPOP | TP53 |
| --- | --- | --- | --- | --- | --- | --- | --- | --- | --- | --- | --- | --- | --- | --- | --- | --- |
| AR | 0 |  |  |  |  |  |  |  |  |  |  |  |  |  |  |  |
| BRCA2 | 0.19262568 | 0 |  |  |  |  |  |  |  |  |  |  |  |  |  |  |
| CCND1 | 0.53047812 | 1.1186E-05 | 0 |  |  |  |  |  |  |  |  |  |  |  |  |  |
| CDK12 | 0.32130028 | 1.4105E-07 | 0.997367 | 0 |  |  |  |  |  |  |  |  |  |  |  |  |
| FGF19 | 0.65957867 | 0.0001069 | 1 | 0.96913694 | 0 |  |  |  |  |  |  |  |  |  |  |  |
| FGF3 | 0.70057248 | 0.00021456 | 1 | 0.96416129 | 1 | 0 |  |  |  |  |  |  |  |  |  |  |
| FGF4 | 0.6903223 | 0.00018026 | 1 | 0.92615916 | 1 | 1 | 0 |  |  |  |  |  |  |  |  |  |
| FGFR1 | 0.42601094 | 0.0048978 | 0.4682276 | 0.01340237 | 0.6043079 | 0.64890865 | 0.63769272 | 0 |  |  |  |  |  |  |  |  |
| LYN | 0.11578071 | 2.3638E-07 | 0.11336638 | 5.0021E-07 | 0.14538918 | 0.19064472 | 0.17836613 | 0.99998996 | 0 |  |  |  |  |  |  |  |
| MYC | 0.08032441 | 0.00016414 | 0.5894616 | 0.02546144 | 0.68110831 | 0.67490702 | 0.65129187 | 0.99998802 | 1 | 0 |  |  |  |  |  |  |
| PRKCI | 0.48397541 | 0.51790039 | 0.87061017 | 0.51928396 | 0.83995761 | 0.86717055 | 0.86054596 | 0.44128018 | 0.04098228 | 0.30385534 | 0 |  |  |  |  |  |
| PTEN | 7.7329E-06 | 1.2093E-12 | 3.8055E-09 | 3.8738E-21 | 2.4764E-07 | 1.3538E-06 | 8.8859E-07 | 0.00054866 | 6.4751E-09 | 8.899E-14 | 0.08686946 | 0 |  |  |  |  |
| RAD21 | 0.00859618 | 0.05428348 | 0.25044823 | 8.4742E-08 | 0.27114665 | 0.36889797 | 0.25001063 | 0.99973172 | 1 | 1 | 0.13843312 | 1.1602E-18 | 0 |  |  |  |
| RB1 | 0.09826633 | 0.26072846 | 0.0148927 | 0.00021062 | 0.04098044 | 0.05564557 | 0.05156142 | 0.59216888 | 0.02983017 | 0.03787844 | 0.02196546 | 0.00307091 | 0.01670548 | 0 |  |  |
| SPOP | 0.00628308 | 1.3624E-11 | 2.1215E-09 | 7.8051E-13 | 1.4435E-07 | 5.2409E-07 | 3.7987E-07 | 0.04737349 | 6.8319E-08 | 8.278E-10 | 0.00239695 | 4.086E-30 | 5.9892E-13 | 1.2848E-08 | 0 |  |
| TP53 | 0.00145609 | 1.1178E-11 | 3.7748E-06 | 6.8335E-21 | 4.1392E-05 | 8.0834E-05 | 0.00013258 | 0.50829933 | 0.79206866 | 0.05193523 | 0.15339053 | 1.5153E-11 | 0.0853454 | 0.02663225 | 8.2484E-40 | 0 |

**Supplementary Table 3:** Performance comparison of different genetic selection methods.

|  |  |  |  |  |
| --- | --- | --- | --- | --- |
| Method | Features | C-index | Brier | Summary AUC |
| **Cox** | baseline | 0.730 | 0.134 | 0.810 |
|  | baseline + individual genes | Singular Matrix |  |  |
|  | **baseline + genomic classification** | **0.761** | **0.130** | **0.832** |
| Elastic Net Cox | baseline | 0.731 | 0.134 | 0.811 |
|  | baseline + individual genes | 0.668 | 0.165 | 0.723 |
|  | baseline + genomic classification | 0.761 | 0.130 | 0.832 |
| Random survival forest | baseline | 0.713 | 0.150 | 0.770 |
|  | baseline + individual genes | 0.759 | 0.144 | 0.802 |
|  | baseline + genomic classification | 0.746 | 0.143 | 0.800 |
| Gradient Boosting | baseline | 0.733 | 0.138 | 0.806 |
|  | baseline + individual genes | 0.765 | 0.134 | 0.827 |
|  | baseline + genomic classification | 0.761 | 0.134 | 0.823 |

**Supplementary Table 4.** Patient characteristics of the validation MSKCC IMPACT cohort.

| **Variable** | **MSK-IMPACT**<br>**synchronous mHSPC** |
| --- | --- |
| **Age at metastatic PCa Dx** |  |
| Mean (SD) | 65.5 (9.4) |
| Median (IQR) | 65.6 (59.0-71.8) |
| Min,Max | 36.7,90.0 |
| **Ethnicity** |  |
| Non-Hispanic |  |
| Hispanic | 39 (3.8%) |
| other/unknown | 226 (21.8%) |
| **Race** |  |
| White | 837 (80.6%) |
| Black | 87 (8.4%) |
| other/unknown | 115 (11.1%) |
| **Sample Type** |  |
| Primary | 662 (63.7%) |
| Metastasis | 377 (36.3%) |
| **Gleason Grade Group** |  |
| Grade 1 | 10 (1.0%) |
| Grade 2 or 3 | 179 (17.2%) |
| Grade 4 | 154 (14.8%) |
| Grade 5 | 394 (37.9%) |
| Unknown | 267 (25.7%) |
| **Vital Status** |  |
| death | 296 (28.5) |
| **Months of survival** |  |
| Mean (SD) | 26.1 (18.8) |
| Median (IQR) | 21.8 (11.8-38.1) |
| Min,Max | 0,77.7 |
| **Volume of Disease** |  |
| High Volume | 257 (24.7%) |
| Low Volume | 782 (75.3%) |

**Supplementary Table 5:**
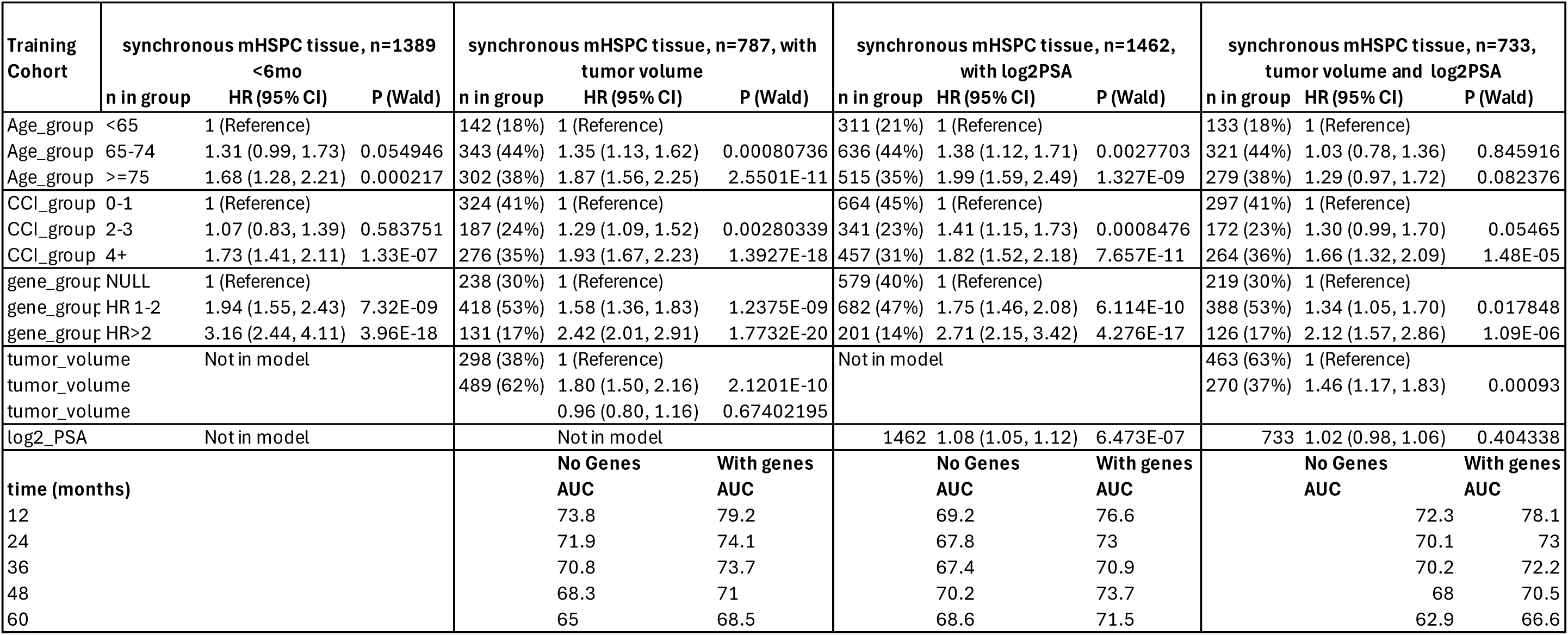
Association of genomic prognostication groups with overall survival in multivariable Cox model with tAUC assessment in patients with known tumor volume and PSA.

**Supplementary Table 6:** Association of genomic prognostication groups with overall survival in multivariable Cox model with tAUC assessment in different disease states and analytes.

|  | Overall Cohort | synchronous mHSPC tissue<br>n=2484 (34% of cohort) | Test synchronous mHSPC tissue |  |  | MSK IMPACT, synchronous mHSPC |  |  | synchronous mHSPC liquid |  |  | metachronous mHSPC tissue |  |  | metachronous mHSPC liquid |  |  |
| --- | --- | --- | --- | --- | --- | --- | --- | --- | --- | --- | --- | --- | --- | --- | --- | --- | --- |
| Level | n=7201<br>n in group | n in group | n in group | n=746 (10% of cohort)<br>HR (95% CI) | P (Wald) | n in group | n=1039<br>HR (95% CI) | P (Wald) | n in group | n=384 (5% of cohort)<br>HR (95% CI) | P (Wald) | n in group | n=1236 (17% of cohort)<br>HR (95% CI) | P (Wald) | n in group | n=297 (4% of cohort)<br>HR (95% CI) | P (Wald) |
| Age_group <65 | 1,290 (18%) | 509 (20%) | 148 (20%) | 1 (Reference) |  | 492 (47%) | 1 (Reference) |  | 60 (16%) | 1 (Reference) |  | 194 (16%) | 1 (Reference) |  | 35 (12%) | 1 (Reference) |  |
| Age_group 65-74 | 3,137 (44%) | 1,040 (42%) | 313 (42%) | 1.54 (1.09, 2.17) | 0.013544 | 382 (37%) | 1.38 (1.05, 1.79) | 0.018641 | 136 (35%) | 1.24 (0.66, 2.36) | 0.502009 | 592 (48%) | 1.16 (0.87, 1.55) | 0.301572 | 107 (36%) | 1.04 (0.39, 2.81) | 0.934073 |
| Age_group >=75 | 2,774 (39%) | 935 (38%) | 285 (38%) | 2.02 (1.41, 2.89) | 0.000123 | 165 (16%) | 2.71 (2.00, 3.68) | 1.37E-10 | 188 (49%) | 1.55 (0.84, 2.86) | 0.16355 | 450 (36%) | 2.33 (1.72, 3.15) | 3.62E-08 | 155 (52%) | 2.41 (0.93, 6.24) | 0.071027 |
| CCI_group 0-1 | 4,393 (61%) | 1,122 (45%) | 334 (45%) | 1 (Reference) |  |  |  |  | 155 (40%) | 1 (Reference) |  | 849 (69%) | 1 (Reference) |  | 236 (79%) | 1 (Reference) |  |
| CCI_group 2-3 | 1,486 (21%) | 609 (25%) | 187 (25%) | 1.05 (0.77, 1.42) | 0.770424 |  |  |  | 100 (26%) | 2.07 (1.24, 3.46) | 0.005308 | 265 (21%) | 1.52 (1.19, 1.93) | 0.000639 | 44 (15%) | 2.28 (1.06, 4.87) | 0.034098 |
| CCI_group 4+ | 1,322 (18%) | 753 (30%) | 225 (30%) | 2.23 (1.71, 2.92) | 4.63E-09 |  |  |  | 129 (34%) | 2.32 (1.44, 3.72) | 0.000524 | 122 (9.9%) | 1.80 (1.33, 2.43) | 0.000132 | 17 (5.7%) | 3.98 (1.50, 10.57) | 0.005673 |
| gene_group NULL | 2,721 (38%) | 947 (38%) | 266 (36%) | 1 (Reference) |  | 457 (44%) | 1 (Reference) |  | 200 (52%) | 1 (Reference) |  | 544 (44%) | 1 (Reference) |  | 174 (59%) | 1 (Reference) |  |
| gene_group HR 1-2 | 3,469 (48%) | 1,180 (48%) | 365 (49%) | 1.77 (1.34, 2.33) | 5.86E-05 | 453 (44%) | 2.45 (1.87, 3.21) | 7.46E-11 | 151 (39%) | 2.23 (1.45, 3.43) | 0.000279 | 556 (45%) | 1.45 (1.17, 1.79) | 0.000725 | 111 (37%) | 2.81 (1.47, 5.36) | 0.001698 |
| gene_group HR>2 | 1,011 (14%) | 357 (14%) | 115 (15%) | 2.81 (2.02, 3.92) | 1.06E-09 | 129 (12%) | 4.37 (3.06, 6.22) | 3.4E-16 | 33 (8.6%) | 5.13 (2.93, 8.97) | 1.03E-08 | 136 (11%) | 2.06 (1.54, 2.76) | 1.24E-06 | 12 (4.0%) | 9.00 (3.14, 25.82) | 4.4E-05 |
| time (months) |  |  |  | AUC |  |  | AUC |  |  | AUC |  |  | AUC |  |  | AUC |  |
| 12 |  |  |  | 76.6 |  |  | 75 |  |  | 70.3 |  |  | 69.5 |  |  | 79.4 |  |
| 24 |  |  |  | 72.1 |  |  | 70.9 |  |  | 76.8 |  |  | 64.5 |  |  | 74.6 |  |
| 36 |  |  |  | 71.1 |  |  | 70.8 |  |  | 74.1 |  |  | 65 |  |  | 78.8 |  |
| 48 |  |  |  | 72 |  |  | 71.8 |  |  | 75.7 |  |  | 62.5 |  |  | 76.7 |  |
| 60 |  |  |  | 69.7 |  |  | 70.9 |  |  | 72.8 |  |  | 63.8 |  |  | 84.4 |  |

| Level | synchronous mCRPC tissue<br>n=336 (5% of cohort) |  |  | synchronous mCRPC liquid<br>n=691 (10% of cohort) |  |  | metachronous mCRPC tissue<br>n=488 (7% of cohort) |  |  | metachronous mCRPC liquid<br>n=1285 (18% of cohort) |  |  |
| --- | --- | --- | --- | --- | --- | --- | --- | --- | --- | --- | --- | --- |
|  | n in group | HR (95% CI) | P (Wald) | n in group | HR (95% CI) | P (Wald) | n in group | HR (95% CI) | P (Wald) | n in group | HR (95% CI) | P (Wald) |
| Age_group <65 | 103 (31%) | 1 (Reference) |  | 178 (26%) | 1 (Reference) |  | 67 (14%) | 1 (Reference) |  | 144 (11%) | 1 (Reference) |  |
| Age_group 65-74 | 144 (43%) | 1.24 (0.91, 1.70) | 0.178375 | 282 (41%) | 1.95 (1.49, 2.56) | 1.23E-06 | 256 (52%) | 1.42 (1.02, 1.97) | 0.03795 | 580 (45%) | 1.38 (1.04, 1.81) | 0.024234 |
| Age_group >=75 | 89 (26%) | 2.37 (1.65, 3.38) | 2.43E-06 | 231 (33%) | 2.83 (2.11, 3.81) | 6.11E-12 | 165 (34%) | 2.59 (1.82, 3.68) | 1.15E-07 | 561 (44%) | 2.81 (2.12, 3.73) | 8.06E-13 |
| CCI_group 0-1 | 188 (56%) | 1 (Reference) |  | 393 (57%) | 1 (Reference) |  | 402 (82%) | 1 (Reference) |  | 1,048 (82%) | 1 (Reference) |  |
| CCI_group 2-3 | 82 (24%) | 1.33 (0.95, 1.87) | 0.093868 | 151 (22%) | 1.37 (1.06, 1.77) | 0.016316 | 69 (14%) | 1.40 (1.01, 1.93) | 0.041116 | 166 (13%) | 1.17 (0.91, 1.50) | 0.217932 |
| CCI_group 4+ | 66 (20%) | 2.19 (1.55, 3.11) | 1.05E-05 | 147 (21%) | 2.05 (1.57, 2.68) | 1.31E-07 | 17 (3.5%) | 1.57 (0.89, 2.78) | 0.118722 | 71 (5.5%) | 1.81 (1.31, 2.51) | 0.000343 |
| gene_group NULL | 60 (18%) | 1 (Reference) |  | 224 (32%) | 1 (Reference) |  | 76 (16%) | 1 (Reference) |  | 496 (39%) | 1 (Reference) |  |
| gene_group HR 1-2 | 185 (55%) | 1.41 (0.95, 2.10) | 0.087151 | 360 (52%) | 2.04 (1.58, 2.63) | 3.8E-08 | 275 (56%) | 1.70 (1.18, 2.44) | 0.004606 | 651 (51%) | 1.93 (1.58, 2.35) | 5.93E-11 |
| gene_group HR>2 | 91 (27%) | 2.03 (1.31, 3.14) | 0.001492 | 107 (15%) | 3.25 (2.39, 4.43) | 8.08E-14 | 137 (28%) | 2.25 (1.53, 3.30) | 3.71E-05 | 138 (11%) | 3.00 (2.31, 3.90) | 2.24E-16 |
| time (months) |  |  | AUC |  |  | AUC |  |  | AUC |  |  | AUC |
| 12 |  |  | 70.4 |  |  | 74.3 |  |  | 70.2 |  |  | 72.3 |
| 24 |  |  | 73.4 |  |  | 70.3 |  |  | 63.1 |  |  | 68.6 |
| 36 |  |  | 70.8 |  |  | 73.9 |  |  | 62.9 |  |  | 68.9 |
| 48 |  |  | 71 |  |  | 76.9 |  |  | 65.7 |  |  | 67.5 |
| 60 |  |  | 69.4 |  |  | 78.4 |  |  | 65.8 |  |  | 68 |

## References

1. Hamid AA, Gray KP, Shaw G, MacConaill LE, Evan C, Bernard B, Loda M, Corcoran NM, Van Allen EM, Choudhury AD, Sweeney CJ: Compound Genomic Alterations of TP53, PTEN, and RB1 Tumor Suppressors in Localized and Metastatic Prostate Cancer. Eur Urol 76:89–97, 2019

2. Attard G, Agarwal N, Graff JN, Sandhu S, Efstathiou E, Ozguroglu M, Pereira de Santana Gomes AJ, Vianna K, Luo H, Gotto GT, Cheng HH, Kim W, Varela CR, Schaeffer D, Kramer K, Li S, Baron B, Shen F, Mundle SD, McCarthy SA, Olmos D, Chi KN, Rathkopf DE: Niraparib and abiraterone acetate plus prednisone for HRR-deficient metastatic castration-sensitive prostate cancer: a randomized phase 3 trial. Nat Med 31:4109–4118, 2025

3. Paik S, Shak S, Tang G, Kim C, Baker J, Cronin M, Baehner FL, Walker MG, Watson D, Park T, Hiller W, Fisher ER, Wickerham DL, Bryant J, Wolmark N: A multigene assay to predict recurrence of tamoxifen-treated, node-negative breast cancer. N Engl J Med 351:2817–26, 2004

4. Marin C, Strawderman M, Peterson D, Cheng Z, Li Y, Gooch JC, Dhakal A, Gergelis KR, Turner B, Brown EB, O’Regan R, Weiss A: Biology is Queen: The Oncotype DX 21-Gene Recurrence Score Has Stronger Prognostic Ability than Lymph Node Burden for Patients with Breast Cancer. Ann Surg Oncol 32:9825–9835, 2025

5. Sparano JA, Crager MR, Tang G, Gray RJ, Stemmer SM, Shak S: Development and Validation of a Tool Integrating the 21-Gene Recurrence Score and Clinical-Pathological Features to Individualize Prognosis and Prediction of Chemotherapy Benefit in Early Breast Cancer. J Clin Oncol 39:557–564, 2021

6. Spratt DE, Zhang J, Santiago-Jimenez M, Dess RT, Davis JW, Den RB, Dicker AP, Kane CJ, Pollack A, Stoyanova R, Abdollah F, Ross AE, Cole A, Uchio E, Randall JM, Nguyen H, Zhao SG, Mehra R, Glass AG, Lam LLC, Chelliserry J, du Plessis M, Choeurng V, Aranes M, Kolisnik T, Margrave J, Alter J, Jordan J, Buerki C, Yousefi K, Haddad Z, Davicioni E, Trabulsi EJ, Loeb S, Tewari A, Carroll PR, Weinmann S, Schaeffer EM, Klein EA, Karnes RJ, Feng FY, Nguyen PL: Development and Validation of a Novel Integrated Clinical-Genomic Risk Group Classification for Localized Prostate Cancer. J Clin Oncol 36:581–590, 2018

7. Brooks MA, Thomas L, Magi-Galluzzi C, Li J, Crager MR, Lu R, Abran J, Aboushwareb T, Klein EA: GPS Assay Association With Long-Term Cancer Outcomes: Twenty-Year Risk of Distant Metastasis and Prostate Cancer-Specific Mortality. JCO Precis Oncol 5:442–449, 2021

8. James ND, Tannock I, N’Dow J, Feng F, Gillessen S, Ali SA, Trujillo B, Al-Lazikani B, Attard G, Bray F, Comperat E, Eeles R, Fatiregun O, Grist E, Halabi S, Haran A, Herchenhorn D, Hofman MS, Jalloh M, Loeb S, MacNair A, Mahal B, Mendes L, Moghul M, Moore C, Morgans A, Morris M, Murphy D, Murthy V, Nguyen PL, Padhani A, Parker C, Rush H, Sculpher M, Soule H, Sydes MR, Tilki D, Tunariu N, Villanti P, Xie LP: The Lancet Commission on prostate cancer: planning for the surge in cases. Lancet 403:1683–1722, 2024

9. Bray F, Laversanne M, Sung H, Ferlay J, Siegel RL, Soerjomataram I, Jemal A: Global cancer statistics 2022: GLOBOCAN estimates of incidence and mortality worldwide for 36 cancers in 185 countries. CA Cancer J Clin 74:229–263, 2024

10. Schoen MW, Montgomery RB, Owens L, Khan S, Sanfilippo KM, Etzioni RB: Survival in Patients With De Novo Metastatic Prostate Cancer. JAMA Netw Open 7:e241970, 2024

11. Elmehrath AO, Afifi AM, Al-Husseini MJ, Saad AM, Wilson N, Shohdy KS, Pilie P, Sonbol MB, Alhalabi O: Causes of Death Among Patients With Metastatic Prostate Cancer in the US From 2000 to 2016. JAMA Netw Open 4:e2119568, 2021

12. D’Amico AV, Whittington R, Malkowicz SB, Schultz D, Blank K, Broderick GA, Tomaszewski JE, Renshaw AA, Kaplan I, Beard CJ, Wein A: Biochemical outcome after radical prostatectomy, external beam radiation therapy, or interstitial radiation therapy for clinically localized prostate cancer. JAMA 280:969–74, 1998

13. Cooperberg MR, Hilton JF, Carroll PR: The CAPRA-S score: A straightforward tool for improved prediction of outcomes after radical prostatectomy. Cancer 117:5039–46, 2011

14. Spratt DE, Tang S, Sun Y, Huang HC, Chen E, Mohamad O, Armstrong AJ, Tward JD, Nguyen PL, Lang JM, Zhang J, Mitani A, Simko JP, DeVries S, van der Wal D, Pinckaers H, Monson JM, Campbell HA, Wallace J, Ferguson MJ, Bahary JP, Schaeffer EM, Sandler HM, Tran PT, Rodgers JP, Esteva A, Yamashita R, Feng FY: Artificial Intelligence Predictive Model for Hormone Therapy Use in Prostate Cancer. NEJM Evid 2:EVIDoa2300023, 2023

15. Parker CTA, Mendes L, Liu VYT, Grist E, Joun S, Yamashita R, Mitani A, Chen E, Parry MA, Sachdeva A, Murphy L, Huang HC, Griffin J, van der Wal D, Todorovic T, Lall S, Santos Vidal S, Goncalves M, Thakali S, Wingate A, Zakka L, Brown M, Wetterskog D, Amos CL, Atako NB, Jones RJ, Cross WR, Gillessen S, Parker CC, collaborators S, Berney DM, Tran PT, Spratt DE, Sydes MR, Parmar MKB, Clarke NW, Brown LC, Feng FY, Esteva A, James ND, Attard G: External validation of a digital pathology-based multimodal artificial intelligence-derived prognostic model in patients with advanced prostate cancer starting long-term androgen deprivation therapy: a post-hoc ancillary biomarker study of four phase 3 randomised controlled trials of the STAMPEDE platform protocol. Lancet Digit Health 7:100885, 2025

16. Halabi S, Yang Q, Roy A, Luo B, Araujo JC, Logothetis C, Sternberg CN, Armstrong AJ, Carducci MA, Chi KN, de Bono JS, Petrylak DP, Fizazi K, Higano CS, Morris MJ, Rathkopf DE, Saad F, Ryan CJ, Small EJ, Kelly WK: External Validation of a Prognostic Model of Overall Survival in Men With Chemotherapy-Naive Metastatic Castration-Resistant Prostate Cancer. J Clin Oncol 41:2736–2746, 2023

17. Sweeney CJ, Chen YH, Carducci M, Liu G, Jarrard DF, Eisenberger M, Wong YN, Hahn N, Kohli M, Cooney MM, Dreicer R, Vogelzang NJ, Picus J, Shevrin D, Hussain M, Garcia JA, DiPaola RS: Chemohormonal Therapy in Metastatic Hormone-Sensitive Prostate Cancer. N Engl J Med 373:737–46, 2015

18. Burdett S, Boeve LM, Ingleby FC, Fisher DJ, Rydzewska LH, Vale CL, van Andel G, Clarke NW, Hulshof MC, James ND, Parker CC, Parmar MK, Sweeney CJ, Sydes MR, Tombal B, Verhagen PC, Tierney JF, Collaborators SMR: Prostate Radiotherapy for Metastatic Hormone-sensitive Prostate Cancer: A STOPCAP Systematic Review and Meta-analysis. Eur Urol 76:115–124, 2019

19. Parker CC, James ND, Brawley CD, Clarke NW, Hoyle AP, Ali A, Ritchie AWS, Attard G, Chowdhury S, Cross W, Dearnaley DP, Gillessen S, Gilson C, Jones RJ, Langley RE, Malik ZI, Mason MD, Matheson D, Millman R, Russell JM, Thalmann GN, Amos CL, Alonzi R, Bahl A, Birtle A, Din O, Douis H, Eswar C, Gale J, Gannon MR, Jonnada S, Khaksar S, Lester JF, O’Sullivan JM, Parikh OA, Pedley ID, Pudney DM, Sheehan DJ, Srihari NN, Tran ATH, Parmar MKB, Sydes MR, Systemic Therapy for Advanced or Metastatic Prostate cancer: Evaluation of Drug Efficacy i: Radiotherapy to the primary tumour for newly diagnosed, metastatic prostate cancer (STAMPEDE): a randomised controlled phase 3 trial. Lancet 392:2353–2366, 2018

20. Grist E, Dutey-Magni P, Parry MA, Mendes L, Sachdeva A, Proudfoot JA, Hamid AA, Ismail M, Howlett S, Friedrich S, DePaula Oliveira L, Murphy L, Brawley C, Dairo O, Lall S, Liu Y, Wetterskog D, Wingate A, Nowakowska K, Zakka L, Amos CL, Atako NB, Wang V, Rush HL, Jones RJ, Leung H, Cross WR, Gillessen S, Parker CC, Marafioti T, Urbanucci A, Fittall M, Schaeffer EM, Spratt DE, Waugh D, Powles T, Sydes MR, Feng FY, Berney DM, Parmar MKB, Clarke NW, Davicioni E, Lotan TL, Sweeney CJ, Brown LC, James ND, Attard G: Tumor transcriptome-wide expression classifiers predict treatment sensitivity in advanced prostate cancers. Cell 188:5717–5734 e10, 2025

21. Robinson D, Van Allen EM, Wu YM, Schultz N, Lonigro RJ, Mosquera JM, Montgomery B, Taplin ME, Pritchard CC, Attard G, Beltran H, Abida W, Bradley RK, Vinson J, Cao X, Vats P, Kunju LP, Hussain M, Feng FY, Tomlins SA, Cooney KA, Smith DC, Brennan C, Siddiqui J, Mehra R, Chen Y, Rathkopf DE, Morris MJ, Solomon SB, Durack JC, Reuter VE, Gopalan A, Gao J, Loda M, Lis RT, Bowden M, Balk SP, Gaviola G, Sougnez C, Gupta M, Yu EY, Mostaghel EA, Cheng HH, Mulcahy H, True LD, Plymate SR, Dvinge H, Ferraldeschi R, Flohr P, Miranda S, Zafeiriou Z, Tunariu N, Mateo J, Perez-Lopez R, Demichelis F, Robinson BD, Schiffman M, Nanus DM, Tagawa ST, Sigaras A, Eng KW, Elemento O, Sboner A, Heath EI, Scher HI, Pienta KJ, Kantoff P, de Bono JS, Rubin MA, Nelson PS, Garraway LA, Sawyers CL, Chinnaiyan AM: Integrative clinical genomics of advanced prostate cancer. Cell 161:1215–1228, 2015

22. Abida W, Cyrta J, Heller G, Prandi D, Armenia J, Coleman I, Cieslik M, Benelli M, Robinson D, Van Allen EM, Sboner A, Fedrizzi T, Mosquera JM, Robinson BD, De Sarkar N, Kunju LP, Tomlins S, Wu YM, Nava Rodrigues D, Loda M, Gopalan A, Reuter VE, Pritchard CC, Mateo J, Bianchini D, Miranda S, Carreira S, Rescigno P, Filipenko J, Vinson J, Montgomery RB, Beltran H, Heath EI, Scher HI, Kantoff PW, Taplin ME, Schultz N, deBono JS, Demichelis F, Nelson PS, Rubin MA, Chinnaiyan AM, Sawyers CL: Genomic correlates of clinical outcome in advanced prostate cancer. Proc Natl Acad Sci U S A 116:11428–11436, 2019

23. Taylor BS, Schultz N, Hieronymus H, Gopalan A, Xiao Y, Carver BS, Arora VK, Kaushik P, Cerami E, Reva B, Antipin Y, Mitsiades N, Landers T, Dolgalev I, Major JE, Wilson M, Socci ND, Lash AE, Heguy A, Eastham JA, Scher HI, Reuter VE, Scardino PT, Sander C, Sawyers CL, Gerald WL: Integrative genomic profiling of human prostate cancer. Cancer Cell 18:11–22, 2010

24. Grasso CS, Wu YM, Robinson DR, Cao X, Dhanasekaran SM, Khan AP, Quist MJ, Jing X, Lonigro RJ, Brenner JC, Asangani IA, Ateeq B, Chun SY, Siddiqui J, Sam L, Anstett M, Mehra R, Prensner JR, Palanisamy N, Ryslik GA, Vandin F, Raphael BJ, Kunju LP, Rhodes DR, Pienta KJ, Chinnaiyan AM, Tomlins SA: The mutational landscape of lethal castration-resistant prostate cancer. Nature 487:239–43, 2012

25. Quigley DA, Dang HX, Zhao SG, Lloyd P, Aggarwal R, Alumkal JJ, Foye A, Kothari V, Perry MD, Bailey AM, Playdle D, Barnard TJ, Zhang L, Zhang J, Youngren JF, Cieslik MP, Parolia A, Beer TM, Thomas G, Chi KN, Gleave M, Lack NA, Zoubeidi A, Reiter RE, Rettig MB, Witte O, Ryan CJ, Fong L, Kim W, Friedlander T, Chou J, Li H, Das R, Li H, Moussavi-Baygi R, Goodarzi H, Gilbert LA, Lara PN, Jr., Evans CP, Goldstein TC, Stuart JM, Tomlins SA, Spratt DE, Cheetham RK, Cheng DT, Farh K, Gehring JS, Hakenberg J, Liao A, Febbo PG, Shon J, Sickler B, Batzoglou S, Knudsen KE, He HH, Huang J, Wyatt AW, Dehm SM, Ashworth A, Chinnaiyan AM, Maher CA, Small EJ, Feng FY: Genomic Hallmarks and Structural Variation in Metastatic Prostate Cancer. Cell 174:758–769 e9, 2018

26. Yu EY, Rumble RB, Agarwal N, Cheng HH, Eggener SE, Bitting RL, Beltran H, Giri VN, Spratt D, Mahal B, Lu K, Crispino T, Trabulsi EJ: Germline and Somatic Genomic Testing for Metastatic Prostate Cancer: ASCO Guideline. J Clin Oncol 43:748–758, 2025

27. Schaeffer EM, Srinivas S, Adra N, An Y, Barocas D, Bitting R, Bryce A, Chapin B, Cheng HH, D’Amico AV, Desai N, Dorff T, Eastham JA, Farrington TA, Gao X, Gupta S, Guzzo T, Ippolito JE, Kuettel MR, Lang JM, Lotan T, McKay RR, Morgan T, Netto G, Pow-Sang JM, Reiter R, Roach M, Robin T, Rosenfeld S, Shabsigh A, Spratt D, Teply BA, Tward J, Valicenti R, Wong JK, Shead DA, Snedeker J, Freedman-Cass DA: Prostate Cancer, Version 4.2023, NCCN Clinical Practice Guidelines in Oncology. J Natl Compr Canc Netw 21:1067–1096, 2023

28. Alba PR, Gao A, Lee KM, Anglin-Foote T, Robison B, Katsoulakis E, Rose BS, Efimova O, Ferraro JP, Patterson OV, Shelton JB, Duvall SL, Lynch JA: Ascertainment of Veterans With Metastatic Prostate Cancer in Electronic Health Records: Demonstrating the Case for Natural Language Processing. JCO Clin Cancer Inform 5:1005–1014, 2021

29. Kelley MJ: VA National Precision Oncology Program. Fed Pract 37:S22–S27, 2020

30. Valle LF, Li J, Desai H, Hausler R, Haroldsen C, Chatwal M, Ojo M, Kelley MJ, Rebbeck TR, Rose BS, Rettig MB, Nickols NG, Garraway IP, Yamoah K, Maxwell KN: Somatic Tumor Next-Generation Sequencing in US Veterans With Metastatic Prostate Cancer. JAMA Netw Open 8:e259119, 2025

31. Candelieri-Surette D, Hung A, Lynch JA, Pridgen KM, Agiri FY, Li W, Aggarwal H, Anglin-Foote T, Lee KM, Perez C, Reed S, DuVall SL, Wong YN, Alba PR: Development and Validation of a Tool to Identify Patients Diagnosed With Castration-Resistant Prostate Cancer. JCO Clin Cancer Inform 7:e2300085, 2023

32. Nguyen B, Fong C, Luthra A, Smith SA, DiNatale RG, Nandakumar S, Walch H, Chatila WK, Madupuri R, Kundra R, Bielski CM, Mastrogiacomo B, Donoghue MTA, Boire A, Chandarlapaty S, Ganesh K, Harding JJ, Iacobuzio-Donahue CA, Razavi P, Reznik E, Rudin CM, Zamarin D, Abida W, Abou-Alfa GK, Aghajanian C, Cercek A, Chi P, Feldman D, Ho AL, Iyer G, Janjigian YY, Morris M, Motzer RJ, O’Reilly EM, Postow MA, Raj NP, Riely GJ, Robson ME, Rosenberg JE, Safonov A, Shoushtari AN, Tap W, Teo MY, Varghese AM, Voss M, Yaeger R, Zauderer MG, Abu-Rustum N, Garcia-Aguilar J, Bochner B, Hakimi A, Jarnagin WR, Jones DR, Molena D, Morris L, Rios-Doria E, Russo P, Singer S, Strong VE, Chakravarty D, Ellenson LH, Gopalan A, Reis-Filho JS, Weigelt B, Ladanyi M, Gonen M, Shah SP, Massague J, Gao J, Zehir A, Berger MF, Solit DB, Bakhoum SF, Sanchez-Vega F, Schultz N: Genomic characterization of metastatic patterns from prospective clinical sequencing of 25,000 patients. Cell 185:563–575 e11, 2022

33. Nguyen B: Genomic characterization of metastatic patterns from prospective clinical sequencing of 25,000 patients. Zenodo., 2022

34. Lenis AT, Ravichandran V, Brown S, Alam SM, Katims A, Truong H, Reisz PA, Vasselman S, Nweji B, Autio KA, Morris MJ, Slovin SF, Rathkopf D, Danila D, Woo S, Vargas HA, Laudone VP, Ehdaie B, Reuter V, Arcila M, Berger MF, Viale A, Scher HI, Schultz N, Gopalan A, Donoghue MTA, Ostrovnaya I, Stopsack KH, Solit DB, Abida W: Microsatellite Instability, Tumor Mutational Burden, and Response to Immune Checkpoint Blockade in Patients with Prostate Cancer. Clin Cancer Res 30:3894–3903, 2024

35. Hiemenz MC, Graf RP, Schiavone K, Harries L, Oxnard GR, Ross JS, Huang RSP: Real-World Comprehensive Genomic Profiling Success Rates in Tissue and Liquid Prostate Carcinoma Specimens. Oncologist 27:e970–e972, 2022

36. Benjamini Y, Hochberg Y: Controlling the False Discovery Rate: A Practical and Powerful Approach to Multiple Testing. Journal of the Royal Statistical Society Series B: Statistical Methodology 57:289–300, 1995

37. Quan H, Sundararajan V, Halfon P, Fong A, Burnand B, Luthi JC, Saunders LD, Beck CA, Feasby TE, Ghali WA: Coding algorithms for defining comorbidities in ICD-9-CM and ICD-10 administrative data. Med Care 43:1130–9, 2005

38. Moon I, Groha S, Gusev A: SurvLatent ODE : A Neural ODE based time-to-event model with competing risks for longitudinal data improves cancer-associated Venous Thromboembolism (VTE) prediction. Presented at the Proceedings of the 7th Machine Learning for Healthcare Conference, Proceedings of Machine Learning Research, 2022

39. Schoen MW, Doherty J, Eaton D, Khan S, Candelieri D, Fedele N, Baxi P, Wynveen M, Pickett C, Wilson RJ, Stackable K, Ingram K, Karunanandaa K, Agarwal R, Rajasekhar A, Riekhof F, Govindan S, Cheranda N, Knoche EM, Etzioni RD, Montgomery RB: Treatment Patterns and Survival Among Veterans With De Novo Metastatic Hormone-Sensitive Prostate Cancer. JAMA Netw Open 8:e259433, 2025

40. Pölsterl S: scikit-survival: A Library for Time-to-Event Analysis Built on Top of scikit-learn. Journal of Machine Learning Research 21:1–6, 2020

41. El-Taji O, Taktak S, Jones C, Brown M, Clarke N, Sachdeva A: Cardiovascular Events and Androgen Receptor Signaling Inhibitors in Advanced Prostate Cancer: A Systematic Review and Meta-Analysis. JAMA Oncol 10:874–884, 2024

